# Protocol and Statistical Analysis Plan for the Waveform Capnography versus Colorimetric Carbon Dioxide Detection During Tracheal Intubation of Critically Ill Adults (WAVE) Study

**DOI:** 10.64898/2026.10.07.26364973

**Authors:** Aaron E. Robinson, Matthew E. Prekker, Avignat S. Patel, Michael Waxman, John P. Gaillard, Shilpa Sridhar, Bjorn Westgard, Akram Khan, J. Taylor Herbert, Alexandra Schick, Kevin G. Buell, Michael Gottlieb, Alec Bunting, Fraser Mackay, Alexander Bracey, Jordan E. Nogle, Matthew C. Exline, Matthew Dillon, Vijay Krishnamoorthy, Karen E. Jackson, Thomas Alcorn, Taya Haas, Ian Ward A. Maia, Wesley H. Self, Todd W. Rice, Matthew W. Semler, Jonathan D. Casey, Lianne Siegel, Joseph S. Koopmeiners, Joanna Kuo, Michael A. Puskarich, Brian E. Driver

## Abstract

**Background:** During emergency tracheal intubation, clinicians use waveform capnography or colorimetric carbon dioxide (CO_2_) detection to confirm exhaled CO_2_ from the endotracheal tube to determine its location. Both devices reliably yield a negative result when the tube is in the esophagus. However, in most emergency intubations the tube is correctly located in the trachea, and in this circumstance the relative diagnostic accuracy of the two devices is uncertain. If one device more often fails to detect exhaled CO_2_ despite correct tube placement (a false negative result), this could lead to unnecessary tube removal, repeated intubation attempts, and an increased risk of serious complications.

**Research Question:** Among critically ill adults undergoing emergency tracheal intubation, is waveform capnography or colorimetric CO_2_ detection more sensitive for identifying an endotracheal tube correctly placed in the trachea?

**Study Design and Methods:** The WAVE study is a pragmatic, prospective, paired diagnostic accuracy study conducted in emergency departments and intensive care units in the United States. The study compares waveform capnography and colorimetric CO_2_ detection (index tests) against a non-CO_2_-based reference standard for endotracheal tube location among 2,092 critically ill adults undergoing tracheal intubation. The primary outcome is the sensitivity of each index test for identifying a tube located in the trachea. The secondary outcome is specificity for identifying a tube located outside the trachea.

**Results:** Enrollment began on April 21, 2025 and is ongoing, with completion anticipated in 2026.

**Interpretation:** The WAVE study will provide prospective evidence comparing waveform capnography with colorimetric CO_2_ detection for determination of endotracheal tube location in critically ill adults. These findings may inform clinical practice and guideline recommendations regarding the optimal test of exhaled CO_2_ for use in the ED and ICU. Prespecifying the protocol and statistical analysis plan before completion of enrollment enhances the rigor, reproducibility, and transparency of the study.

## Introduction

Tracheal intubation is a common and high-risk procedure among critically ill patients in the emergency department (ED) and intensive care unit (ICU). To perform tracheal intubation, clinicians attempt to (i) use a laryngoscope or flexible bronchoscope to view the patient’s vocal cords, (ii) place an endotracheal tube between the vocal cords and into the trachea, and (iii) verify the location of the endotracheal tube. Determining whether the endotracheal tube is located in the trachea is critically important because an endotracheal tube located in the esophagus cannot ventilate the lungs. If unrecognized, the sedated and paralyzed patient may rapidly experience severe hypoxemia, brain damage, and death.^1^

Historically, clinicians attempted to determine the location of the endotracheal tube using physical examination alone. While ventilating the patient through the endotracheal tube, clinicians would auscultate for bilateral breath sounds, observe for chest rise, and look for condensation inside the endotracheal tube. It is now recognized that physical examination alone is insufficiently accurate to determine the location of the endotracheal tube.^2^ Beginning in the late 1980s, physical examination was supplanted by tests that detect carbon dioxide (CO_2_) in the gas exhaled via the endotracheal tube. These tests rely on the fact that the lungs eliminate CO_2_ from the body, while the stomach and esophagus do not. The sustained presence of CO_2_ in exhaled gas demonstrates alveolar ventilation and is used clinically to exclude esophageal placement and usually indicates a tube located in the trachea. In contrast, the absence of sustained CO_2_ after endotracheal tube placement alerts the clinician to a potential problem. In this scenario, the absence of sustained CO_2_ can be a result of a misplaced tube (e.g., located in the esophagus), attenuated CO_2_ delivery from the blood to the lungs (e.g., cardiac arrest or shock), or malfunction of the CO_2_ detection equipment.^3^ If a CO_2_ detection device fails to demonstrate exhaled CO_2_ despite correct placement of the endotracheal tube in the trachea in a critically ill patient (a false negative result), the clinician may remove the tube unnecessarily and interrupt ventilation to re-attempt intubation which increases the risk of hypoxemia, hypotension, aspiration, and cardiac arrest.

Two tests to detect exhaled CO_2_ during intubation are in widespread use in the United States: colorimetric CO_2_ detection and waveform capnography. Both are attached to the endotracheal tube adapter and reside between the tube and the bag-valve device or mechanical ventilator. A colorimetric CO_2_ detector uses pH-sensitive paper that dynamically changes color depending on the presence or absence of CO_2_.^4,5^ The paper appears purple when CO_2_ partial pressure is < 4 mm Hg (as occurs during inhalation), tan when CO_2_ is 4-15 mm Hg, and yellow when CO_2_ is > 15 mm Hg (as occurs during exhalation). Colorimetric CO_2_ detectors are portable, disposable, relatively inexpensive, and provide qualitative information regarding the presence or absence of CO_2_ (but not detailed quantitative information on levels of CO_2_). Waveform capnography continuously samples gas from the tube and, using infrared spectroscopy, displays both the CO_2_ waveform over time (capnography) and a numeric value for the breath-to-breath maximal exhaled CO_2_ (ie, end-tidal CO_2_).^6–7,8^ Waveform capnography components include a disposable adapter and a reusable analytic and display module; it is generally more expensive than colorimetric CO_2_ detection and requires a power source.

Existing studies evaluating the diagnostic accuracy of either waveform capnography or colorimetric CO_2_ detection during emergency intubation have substantial limitations. They were performed decades ago among a relatively small number of patients, were conducted at single centers, used CO_2_ detectors that are no longer clinically available, often evaluated patients in the operating room or prehospital setting rather than in the ED or ICU, and used clinical criteria such as auscultation or chest radiography—methods since shown to be unreliable—as the reference standard for endotracheal tube location.^9^ In this context, these studies reported that waveform capnography and colorimetric CO_2_ detection reliably indicated that exhaled CO_2_ was absent when the endotracheal tube was located in the esophagus (i.e., the specificity approached 100%, thereby minimizing “false positive” test results). The ability of these tests to reliably indicate that exhaled CO_2_ was present when the endotracheal tube was correctly located in the trachea was more variable, especially with colorimetric CO_2_ detection use and among patients with cardiac arrest receiving CPR (sensitivity 62-95%, more frequent “false negative” test results). False negative results in these settings could lead clinicians to remove a correctly placed endotracheal tube and repeat the intubation procedure.

In the absence of high quality, contemporary evidence, both waveform capnography and colorimetric CO_2_ detection are in widespread use in EDs and ICUs. In a recent survey of 36 U.S. academic EDs and ICUs, 11% of sites exclusively used waveform capnography, 42% exclusively used colorimetric CO_2_ detection, and the remaining 47% used both tests interchangeably.^10^ In a recent randomized trial of video laryngoscopy in 17 EDs and ICUs in the US, 71% of sites determined tube location with a colorimetric CO_2_ device.^11^ An international, prospective, observational study of 2,964 ED and ICU patients undergoing tracheal intubation found that waveform capnography was the first method used to determine tube location in 25.6% of patients and colorimetric CO_2_ detection was used first in 7.5% of patients (lung auscultation was used first in 57.9% of intubations).^12^

Practice guidelines are also inconsistent or nonspecific regarding the role of colorimetric CO_2_ detection during emergency tracheal intubation. Guidelines by the Project for Universal Management of Airways and the Canadian Airway Focus Group recommended the use of waveform capnography over colorimetric CO_2_ detection for all tracheal intubations^1,13^, but other recent guidelines recommend waveform capnography without specific mention of the role of colorimetric CO_2_ detection^14–16^, and one guideline recommends either “capnography or end tidal carbon dioxide monitoring.”^17^

To address this knowledge gap, the Waveform Capnography vs Colorimetric Carbon Dioxide Detection During Tracheal Intubation of Critically Ill Adults (WAVE) study was designed to compare the performance of waveform capnography versus colorimetric CO_2_ detection for determining the location of the endotracheal tube among critically ill adults undergoing tracheal intubation in the ED or ICU. Because clinicians are trained to remove an endotracheal tube when exhaled CO_2_ is not detected, a device with greater sensitivity for detecting exhaled CO_2_ (i.e. a lower false negative rate) could reduce unnecessary removal of correctly placed tubes and the risks associated with repeated intubation attempts. We hypothesize that waveform capnography has a higher sensitivity than colorimetric CO_2_ detection for detecting exhaled CO_2_ from an endotracheal tube that is correctly located in the trachea.

## Study Design and Methods

This article was written in accordance with STandards for Reporting Diagnostic Accuracy Studies (STARD 2015) (Table 1 and e-Appendix, section 2). This article prespecifies the statistical analyses for the WAVE study before completion of enrollment or unblinding of study data. The WAVE study is being conducted without external funding.

**Table 1.** Schedule of Enrollment, Measurements, and Assessments in the WAVE Study.

| Time Point | Eligibility Screening: Decision to Perform Tracheal Intubation | Before the Tracheal Intubation Procedure | End of the Tracheal Intubation Procedure (0 mins) | After Tracheal Intubation (Up to 20 mins) | Mortality Assessment (28 d) |
| --- | --- | --- | --- | --- | --- |
| **Enrollment** |  |  |  |  |  |
| Eligibility screen | X |  |  |  |  |
| Enrollment | X |  |  |  |  |
| **Interventions** |  |  |  |  |  |
| Application of waveform capnography |  | X |  |  |  |
| Application of colorimetric detector |  | X |  |  |  |
| **Assessments** |  |  |  |  |  |
| Periprocedural vital signs |  | X | X | X |  |
| Interpretation of CO<sub>2</sub> detection devices |  |  | X |  |  |
| Reference standard |  |  |  | X |  |
| Adverse events |  |  |  | X | X |
| Mortality |  |  |  |  | X |

Table entries without data mean that data elements were not collected at that time.

### Patient and public involvement

Study authors will disseminate the results of this study online and via social media in forms suitable for public understanding.

### Study Design

The WAVE study is a pragmatic, prospective, investigator-initiated, paired, comparative diagnostic accuracy study conducted at 5 EDs and 7 ICUs in the United States (additional details in the e-Appendix, section 3). During intubation, both index diagnostic tests—waveform capnography and colorimetric CO_2_ detection—are performed in each patient. The results of these tests are compared with a reference standard that determines the location of the endotracheal tube and does not rely on detection of exhaled CO_2_. The primary outcome is a comparison of the sensitivity of each index test, against the reference standard, for identifying a tube located in the trachea. The secondary outcome is a comparison of the specificity of each index test, against the reference standard, for identifying a tube located outside the trachea. The study is being conducted by the Pragmatic Critical Care Research Group (https://www.pragmaticcriticalcare.org/). The study was registered before initiation of enrollment (ClinicalTrials.gov Identifier: NCT06934876).

### Ethics and Regulatory Approval

The WAVE study protocol was approved by the institutional review board at each participating site. The coordinating center (Hennepin County Medical Center) IRB approved this study. The study is being conducted with a waiver of informed consent, as specified in 45 CFR 46.116(f), as it was deemed to involve minimal incremental risk to patients and obtaining informed consent for participation would be impracticable (additional details in the e-Appendix, section 4).

### Study Population

Critically ill patients in participating EDs and ICUs are eligible for enrollment if they meet all of the inclusion criteria and none of the exclusion criteria.

#### Inclusion criteria

1. The patient is located in a participating unit.
2. The patient is undergoing tracheal intubation.
3. The clinician intends to use an exhaled CO_2_ detection device to confirm tracheal location of an endotracheal tube.

#### Exclusion criteria

1. The patient is known to be a prisoner.
2. The patient is known to be less than 18 years old.
3. A responsible clinician has determined that sole use of either waveform capnography or colorimetric testing is required for optimal care of the patient.

### Enrollment procedures

Patients undergoing tracheal intubation in participating units are screened for eligibility. Before enrollment, the responsible clinical or research personnel perform a “time out” to confirm study eligibility by verbalizing that the patient is 18 years or older, the patient is not a prisoner or in the custody of law enforcement, and that using both waveform capnography and a colorimetric device would be acceptable. Before opening the study envelope containing the data collection sheet, the initials of the clinician who will interpret waveform capnography and the clinician who will interpret the colorimetric CO_2_ detector are recorded on the exterior of the envelope. These assignments help each clinician know ahead of time which device they will interpret, so that they view only that device following intubation. Opening the study envelope signifies enrollment in the study.

### Study Interventions

For all enrolled patients, both the waveform capnography adapter and a colorimetric CO_2_ detection device are attached in series between the endotracheal tube and the device being used to deliver oxygen and ventilation during intubation (e.g., a bag-valve device or mechanical ventilator). The order in which the waveform capnography adapter and colorimetric CO_2_ detection device are attached to the endotracheal tube does not affect the interpretability of either device, so is left up to the treating clinician. The process of interpreting each device is described in the following section. All other aspects of the intubation procedure are determined by the treating clinician.

### Data Collection

A trained observer, not directly involved with the intubation procedure, collects data for key variables before, during, and after the tracheal intubation procedure. These include vital signs, clinicians’ interpretations of waveform capnography and the colorimetric CO_2_ detector (the index tests), and the non-CO_2_-based reference standard to determine endotracheal tube location. The data collection form is shown in section 5 of the e-Appendix.

Information regarding the location of the endotracheal tube (in the trachea vs outside of the trachea) will be obtained from two index tests (waveform capnography and colorimetric CO_2_ detection) and one reference standard, as follows:

### Index Test Interpretation and Data Collection

Two clinicians who normally participate in the tracheal intubation procedure (eg, physicians, advanced practice providers, respiratory therapists) are assigned to interpret the index tests independently, with one clinician assigned to interpret waveform capnography and the other clinician assigned to interpret the colorimetric CO_2_ detector.

Following intubation, the clinician responsible for interpreting waveform capnography views and interprets only that device while the initial 7 or more ventilations are delivered. After at least 7 consecutive breaths are delivered through the endotracheal tube, the clinician responds to the observer’s question: “Is the waveform consistent with a tube located in the trachea?” Simultaneously, the clinician responsible for interpreting the colorimetric CO_2_ detector views and interprets only that device while the initial 7 or more ventilations are delivered and then responds to the observer’s question: “Is the color change consistent with a tube located in the trachea?” Responses are recorded as “yes” (an index-test interpretation consistent with a tracheal location) or “no” (an index-test interpretation not consistent with a tracheal location).

Recognizing the possibility that the first clinician’s verbal interpretation could influence the response of the second clinician, and to prevent one index test from preferentially being interpreted first, we randomly varied the order in which the interpretations of the index tests were solicited by the independent observer. This was accomplished by creating two versions of the data collection form that were identical except for the order of index test interpretation (in one form, the waveform capnography interpretation is queried first, and on the other form, colorimetric CO_2_ detection interpretation is queried first). Sequentially-numbered envelopes contained one of the two versions of the data collection form; which version of the form was contained in each envelope was assigned based on a 1:1 randomization generated using permuted blocks of variable size.

## Waveform Capnography

All three of the following criteria, adapted from the Project for the Universal Management of Airways (PUMA) consensus guideline^1^, are to be present for a clinician to interpret waveform capnography as consistent with a tube located in the trachea (Figure 1). Clinicians at each site received training on these criteria before enrollment began, and the data collection form instructed clinicians to answer “yes” (indicating tracheal location) if all of the following criteria were met:

1. The waveform rises during expiration and falls during inspiration.
2. The amplitude is consistent or increasing over 7 breaths and the peak amplitude exceeds 7.5 mm Hg from baseline.
3. The pattern and amplitude are appropriate for the clinical context.

**Figure 1:**
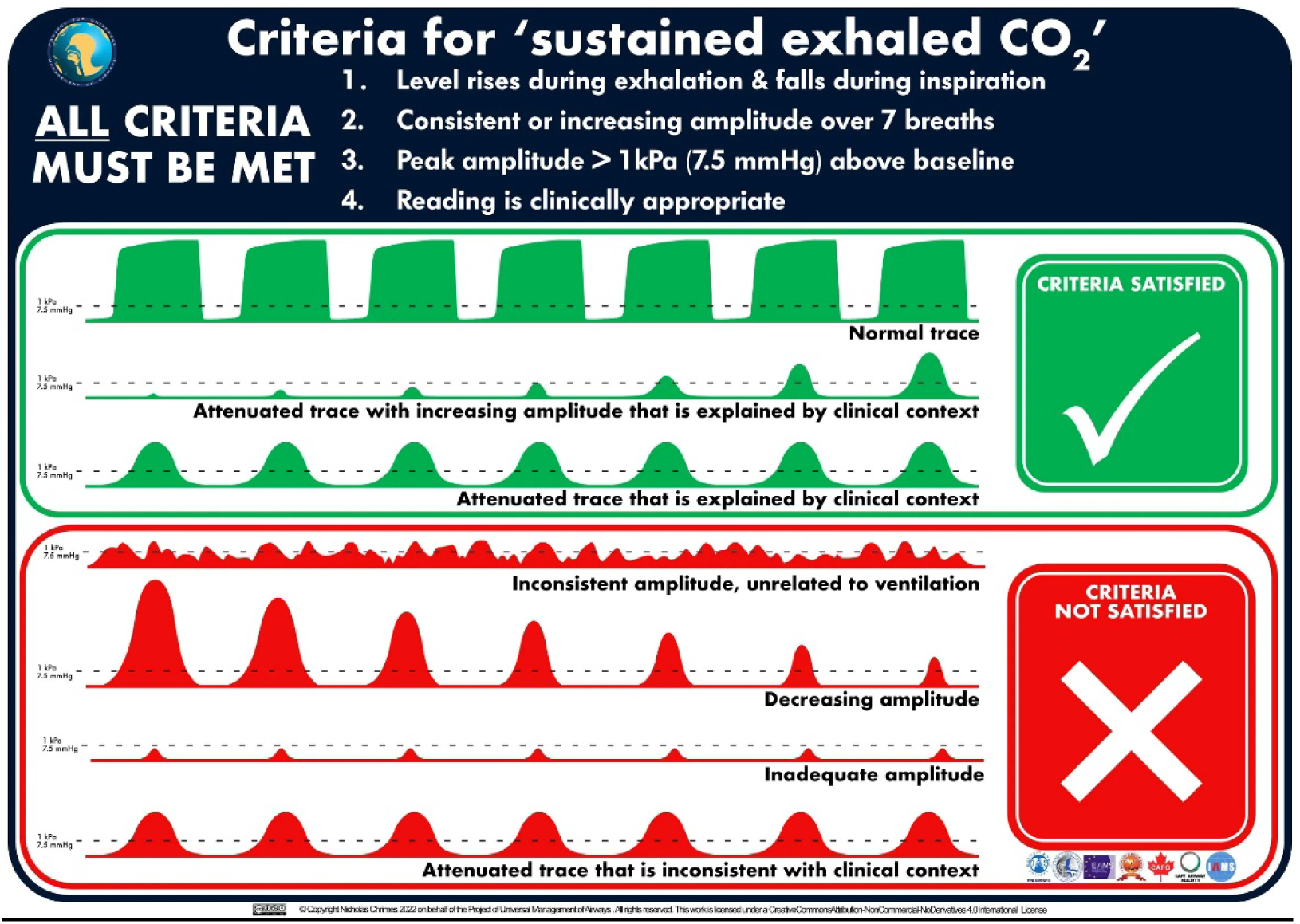
Reproduced with permission under a Creative Commons. Clinicians are trained to interpret the index test as indicating a tube location in the trachea if during at least 7 consecutive breaths, the waveform rises during expiration and falls during inspiration, the amplitude is consistent or increasing over 7 breaths and the peak amplitude exceeds 7.5 mm Hg from baseline, and the pattern and amplitude are appropriate for the clinical context. If the waveform does not meet these criteria, the clinicians are to answer “No” to the question “Is the waveform consistent with a tube located in the trachea?” These criteria are additionally present on every data collection form. Reproduced with permission under a Creative Commons Attribution–Non Commercial–No Derivatives 4.0 International License. © Nicholas Chrimes, Project for Universal Management of Airways, 2022.

If these criteria are not satisfied, the clinicians are to interpret the waveform as not consistent with a tube located in the trachea.

## Colorimetric CO_2_ Detection

The following criteria are to be present for a clinician to interpret the colorimetric CO_2_ detector as consistent with a tube located in the trachea. These criteria were adapted from manufacturer specifications.^18,19^ Clinicians at each site received training on these criteria before enrollment began, and the data collection form instructed clinicians to answer “yes” (indicating tracheal location) if the following criteria were met.

1. During at least 7 consecutive breaths, the color changes to yellow during expiration and returns to purple during inspiration (Figure 2).
2. If the color is tan (between purple and yellow) at the seventh breath, the clinician will deliver 6 more breaths and evaluate for color change. A color remaining tan or changing to yellow with expiration during these additional ventilations will indicate tracheal location of the tube.

**Figure 2:**
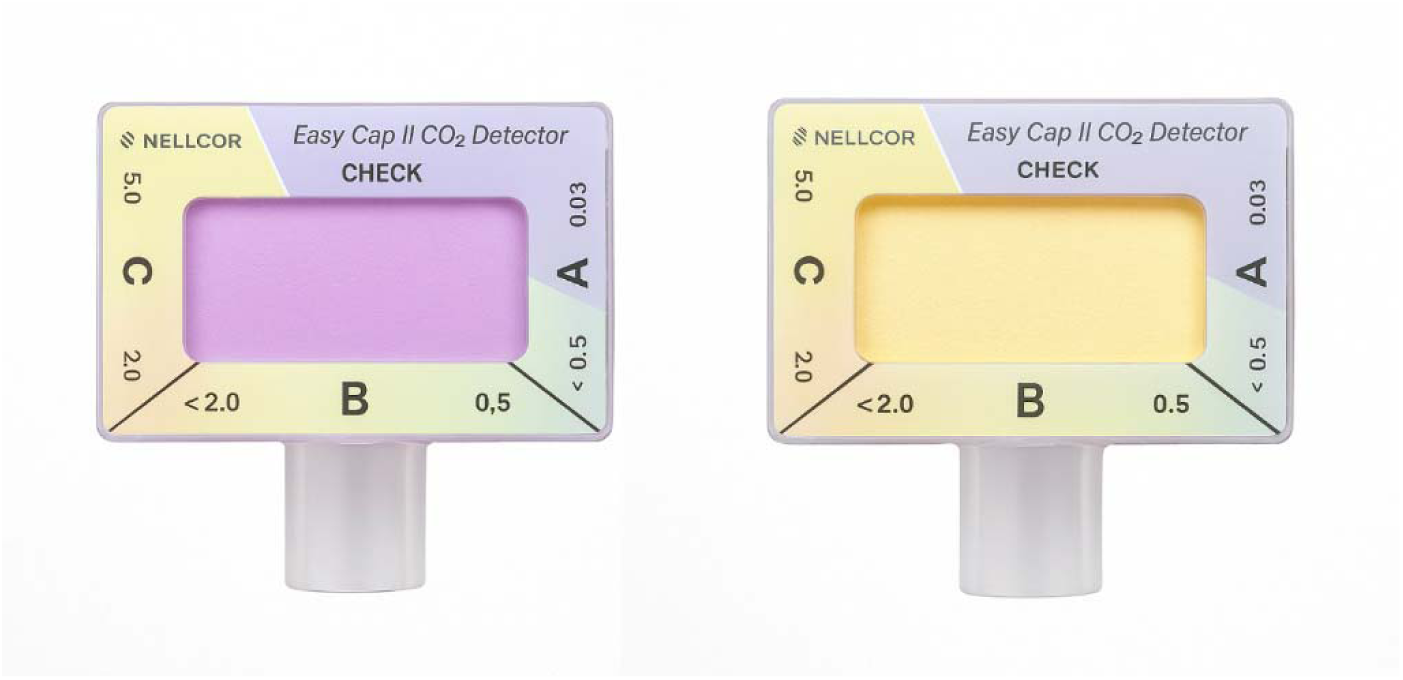
Interpretation of the colorimetric detector index test. Clinicians are trained to interpret the index test as indicating a tube location in the trachea if during at least 7 consecutive breaths, the color changes to yellow during expiration and returns to purple during inspiration. If the color is tan (between purple and yellow) at the seventh breath, the clinician will deliver 6 more breaths and evaluate for color change. A color changing to tan or to yellow with expiration during these additional ventilations will indicate tracheal location of the tube. If the colorimetric CO_2_ detector does not meet these criteria, the clinicians are to answer “No” to the question “Is the color change consistent with a tube located in the trachea?” These criteria are additionally present on every data collection form.

If these criteria are not satisfied, the clinicians are to interpret the colorimetric CO_2_ detector as not consistent with a tube located in the trachea.

## Definition of a Missing Index Test

For the purposes of this study, an index test (waveform capnography or colorimetric CO_2_ detection) is classified as “missing” only if the modality was not used during the intubation procedure. Whenever a modality is used, the clinician must make a definitive determination of ‘yes’ or ‘no’ for tracheal location of the tube using the criteria outlined above. Missing tests will occasionally happen because of logistical issues or from unanticipated clinical urgency.

### Reference Standard Interpretation and Data Collection

The reference standard comprises a structured assessment by the independent observer that was designed to establish the location of the endotracheal tube without relying on exhaled CO_2_. In this pragmatic trial, ‘tracheal placement’ is operationally defined as either direct visual confirmation of the tube within the trachea or objective evidence of sustained, effective gas exchange and ventilation (excluding esophageal placement). The reference standard in this study has three possible values: (1) tracheal, (2) non-tracheal, or (3) indeterminate. The reference standard is recorded by an independent observer not involved in the performance of the tracheal intubation procedure or the interpretation of the index tests of exhaled CO_2_. The independent observer is either trained study personnel (e.g., a research coordinator) or trained clinical personnel (e.g., a trained physician, nurse, or respiratory therapist).

The independent observer’s structured assessment considers the endotracheal tube to be located in the trachea at the first time that *any* of the following criteria have been met: (1) the tube has been connected to a mechanical ventilator in a volume-targeted mode, and the exhaled tidal volume is ≥80% of the delivered tidal volume for five consecutive breaths; (2) the tube is visualized within the trachea during flexible bronchoscopy; or (3) the patient remains intubated with the original tube and maintains spontaneous circulation 20 minutes after intubation. Most critically ill adults undergoing intubation will meet the reference standard by the first criterion assessing exhaled volumes on mechanical ventilation, with the second and third criteria recorded only in cases when the first criterion is not met. In rare cases in which none of these three criteria are satisfied, tracheal location may also be established through consensus review of the video laryngoscopy recording of the index intubation, if available, by two independent clinicians not involved in the intubation procedure who are blinded to the interpretations of the index tests.

The independent observer’s structured assessment considers the endotracheal tube to be located outside the trachea if the endotracheal tube is observed to terminate outside of the trachea on (1) flexible bronchoscopy performed in real-time, or (2) video laryngoscopy performed in real-time or by review of available recordings by independent clinicians.

The independent observer’s structured assessment considers the endotracheal tube location to be indeterminate when the location of the tube cannot be verified to be either in the trachea or outside of the trachea. For example, this may occur if the patient is never connected to a ventilator and the tube is removed, or cardiac arrest or death occur within 20 min—and bronchoscopy is not performed and video laryngoscopy recordings are not available. These indeterminate cases will be excluded from primary diagnostic accuracy analyses but will be accounted for in sensitivity analyses (additional detail for the reference standard is available in Section 6 of the e-Appendix).

This reference standard provides a practical, non-CO_2_-based method for classifying tube location across diverse clinical scenarios. Similar delivered and exhaled tidal volumes on the ventilator confirm functional tracheal ventilation that is not possible if the tube were located in the esophagus.^20–22^ In a human study, exhaled volumes from a tube intentionally placed in the esophagus were typically <10% of the delivered tidal volume, and even with a simulated incompetent gastroesophageal sphincter they did not exceed ∼40% of delivered volume.^20^ Furthermore, experimental animal work demonstrates that, even with prolonged esophageal and gastric ventilation, the esophagus does not behave as a reservoir capable of generating near-normal expired volumes.^21,22^ Bronchoscopy offers direct visualization to confirm tube position without relying on physiologic adjuncts and has served as the sole reference standard in prior airway studies.^2,19,23^ Persistent spontaneous circulation ≥ 20 minutes after intubation also effectively excludes esophageal placement: even in healthy patients with optimal preoxygenation, critical hypoxemia occurs during the first 10 min of apnea,^24^ and in critically ill patients often within 1 min.^25^ Moreover, reported cases of unrecognized esophageal intubation uniformly progress to cardiac arrest within 1–15 minutes.^1,26–28^ Finally, the independent, blinded video laryngoscopy review ensures that cases lacking physiologic or bronchoscopic confirmation are adjudicated in a standardized, reproducible manner when video review is available.

In most cases the reference standard assessment will be completed by the independent observer using the mechanical ventilation criterion in less than 30 seconds (eg, 5 ventilations delivered at a rate of 12 per min would require 25 seconds to assess the exhaled tidal volume). However, the reference standard assessment may be completed by the independent observer up to 20 minutes after intubation, which distinguishes it from the two index tests using CO_2_ detection over the initial 7 breaths delivered through the index endotracheal tube. This potential delay of up to 20 minutes in reference standard completion will not affect the internal validity of this study; in fact, this potential delay is analogous to accepted practice in oncology studies evaluating the diagnostic accuracy of index tests in determining if a lung nodule is benign or malignant which use cancer diagnosis by 1 year as the reference standard.^29,30^

### Blinding

Complete blinding is not possible in this paired, diagnostic test accuracy study. Both index tests were interpreted by clinicians participating in the patient’s care, with access to clinical information ordinarily available during emergency intubation, including observations made during laryngoscopy. This approach preserves the intended clinical context of testing and is consistent with the principle articulated in QUADAS guidance that clinical information available during diagnostic-test evaluation should reflect that available in practice.^31^

Because both index tests require in-line sampling of exhaled gas, the waveform capnography CO_2_ sensor and the colorimetric CO_2_ detector must be attached to the endotracheal tube adapter in close physical proximity, and the two clinicians interpreting them are necessarily present together at the bedside. A clinician assigned to one index test could therefore, in principle, view the other device or overhear the other clinician’s interpretation.

We implemented several measures to limit the resulting potential for bias. First, the index tests are interpreted before the reference standard is known. Second, each index test was assigned to a specific clinician before the procedure began, so they could focus on interpreting that particular index test. Third, each site used waveform capnography that displayed on a separate vital sign monitor and a colorimetric CO_2_ detector that was attached in-line with the endotracheal tube; this allowed the two clinicians to be looking in different locations when interpreting the tests. Fourth, each index test was interpreted independently: to limit the possibility that hearing the first interpretation could influence the second, the two clinicians were instructed to view only their assigned device and to commit to their interpretation before the interpretation for either modality was solicited. Fifth, to prevent one test from consistently being interpreted first, the order in which the independent observer solicited the two interpretations was randomized 1:1 using the two versions of the data collection form described above. Sixth, both index tests are interpreted against explicit, prespecified criteria (Figures 1 and 2) on which clinicians were trained before enrollment, and which were shown on the data collection form, reducing the latitude for a subjective judgment to be swayed by any knowledge of the other result.

A lack of blinding could also theoretically affect determination of the reference standard. The independent observer necessarily hears the index-test interpretations and is therefore not blinded to them; however, the reference standard is established after the index tests using objective criteria, including exhaled tidal volume on the ventilator, bronchoscopic visualization, or maintenance of spontaneous circulation for 20 minutes without tube removal, that cannot be altered by the observer’s awareness of the index-test results. The one component that involves a higher degree of interpretation, the video-laryngoscopy adjudication used in the small number of otherwise unresolved cases, is performed by clinicians blinded to the index-test interpretations.

### Post-Procedure data collection

In the 20 minutes following intubation, the observer records the reference standard, results of any bronchoscopy or repeat laryngoscopy, and any removal of the original tube and reason for removal. The post-procedure systolic blood pressure and first end-tidal CO_2_ value from waveform capnography are also recorded.

Following intubation, the clinician involved with the procedure uses a structured data collection form to record patient characteristics and procedural details including whether troubleshooting either index test was required before interpretation. Next, the clinicians who interpreted the index tests each estimate the number of times they have previously used waveform capnography and colorimetric CO_2_ detection to determine the location of an endotracheal tube and record those estimates on the form.

During the index hospitalization, study personnel at each site review the electronic health record to collect additional data on baseline patient characteristics, co-morbid conditions, active medical problems, management before and after laryngoscopy, route and outcomes of tracheal intubation, and in-hospital mortality by 28 days.

### Primary Outcome

The primary outcome is the sensitivity of the index test, defined as the probability that the test indicates tracheal placement conditional on the reference standard meeting criteria for tracheal placement. A rationale for the primary outcome can be found in section 7 of e-Appendix 1. Briefly, when an endotracheal tube is correctly located in the trachea but an index test fails to demonstrate exhaled CO_2_ (a false negative result), clinicians may interpret this as evidence of esophageal intubation and remove the tube. Such unnecessary removal of a correctly placed endotracheal tube may expose critically ill patients to additional intubation attempts and increased risks of hypoxemia, hypotension, aspiration, and cardiac arrest. Therefore, a diagnostic test with higher sensitivity for detecting tracheal tube placement could reduce unnecessary tube removal and the risks associated with repeated attempts at intubation.

### Secondary Outcome

The secondary outcome is the specificity of the index test, defined as the probability that the test indicates non-tracheal placement conditional on the reference standard meeting criteria for non-tracheal placement. A test with high specificity would correctly identify an endotracheal tube as not located in the trachea when the tube is truly outside the trachea. Prior studies suggest that both waveform capnography and colorimetric CO detection have near-perfect specificity for identifying esophageal intubation; therefore specificity is included as a secondary outcome to provide a complete assessment of diagnostic accuracy. A more detailed rationale for the secondary outcome can be found in section 8 of e-Appendix 1.

### Exploratory Outcomes

We will report exploratory diagnostic accuracy outcomes including the positive predictive value, the negative predictive value, the positive likelihood ratio, and the negative likelihood ratio for each index test to facilitate clinical interpretation. These measures will be presented descriptively without hypothesis testing.

We will also report exploratory outcomes related to troubleshooting of each index test. Troubleshooting is defined as any additional action required to obtain interpretable test output after initial application of the device to the endotracheal tube. Troubleshooting is reported by a clinician after the procedure on the data collection form. Examples include repositioning the device, cleaning the device, replacement of the device, replacement or adjustment of the monitoring module, and changing monitor settings. For each index test, we will report the proportion of cases requiring troubleshooting, the proportion in which troubleshooting successfully resulted in interpretable output, and among cases where troubleshooting resulted in interpretable output, the duration of troubleshooting. Troubleshooting outcomes are not measures of diagnostic accuracy and will be interpreted descriptively to characterize the practical usability and operational burden of each modality in real-world clinical settings.

### Data and Safety Monitoring Board

The data and safety monitoring board (DSMB) approved a charter and study monitoring plan. The DSMB has the authority to recommend that the study stop at any point, to request additional data, to request additional interim analyses, or to request modifications to the study protocol. See section 9 of the e-Appendix for additional details regarding the composition and responsibilities of the DSMB.

### Statistical Analysis Plan and Interim Analysis

#### Statistical Analysis Principles

This statistical analysis plan was finalized and publicly posted prior to completion of enrollment and unblinded analysis of study data. All analyses described herein are considered pre-specified *a priori* analyses.

Analyses will be conducted following reproducible research principles. Categorical variables will be presented as number and percentage. Continuous variables will be presented as mean (SD) or median (interquartile range). Statistical tests will be completed with an overall type-I error rate of 0.05. With a single primary outcome, no adjustment for multiplicity will be made. For other analyses, emphasis will be placed on the magnitude of differences between groups with 95% confidence intervals, rather than statistical significance. P values will not be presented for secondary and exploratory analyses. Exact and score-based methods will be used to ensure valid inference and confidence intervals in the setting of very high sensitivities and potentially small numbers of discordant paired observations.

#### Main Analysis of the Primary Outcome

The main analysis will compare the sensitivity of waveform capnography with that of colorimetric CO_2_ detection for identifying a tube located in the trachea, as determined by the reference standard, among all patients in whom both index tests were interpreted. Sensitivity will be calculated as the number of patients for whom both the index test and the reference standard indicate that the endotracheal tube is in the trachea (true positives) divided by the number of patients for whom the reference standard indicates that the endotracheal tube is in the trachea regardless of the results of the index test (true positives + false negatives), with 95% exact binomial confidence intervals (Clopper-Pearson). Consistent with recommendations for paired diagnostic accuracy studies,^32^ the sensitivity of the two index tests will be compared using McNemar’s exact test within the subset of patients with a reference standard indicating a tube located in the trachea. McNemar’s test is well suited to paired data in which both diagnostic tests are applied to the same patient. The relative performance of the two index tests will be summarized by the absolute difference in sensitivity with 95% score-based confidence intervals for paired proportions (Newcombe method).

#### Sample Size Determination

The original protocol specified a total enrollment of 1,332 patients, which provided 95% power to detect a 2-percentage-point absolute difference in sensitivity between waveform capnography and colorimetric CO_2_ detection for identifying a tube located in the trachea, assuming 99% sensitivity for waveform capnography, 97% sensitivity for colorimetry, and allowing for 3% incomplete data (section 10 of the e-Appendix). At 50% enrollment, a sample size re-estimation was conducted as detailed below in *Sample Size Re-estimation*.

#### Interim Analysis

On December 17, 2025, the DSMB reviewed a single interim analysis, prepared by an unblinded study statistician at the halfway point of the trial after enrollment of 666 patients. The pre-specified Haybittle-Peto stopping boundary for efficacy was a P value < 0.001 for the difference in sensitivity between waveform capnography and colorimetric CO_2_ detection, evaluated using McNemar’s exact test for paired data among patients meeting the reference standard for tracheal tube placement. This conservative stopping boundary limits early spending of type I error, allowing the final analysis to use a two-sided α=0.049. After reviewing the interim analysis, the DSMB recommended continuing the WAVE trial without modification to study conduct, procedures, outcomes, or safety monitoring.

#### Sample Size Re-estimation

During routine review of study data collection forms for completeness and accuracy, investigators observed that the sensitivity of the index tests appeared qualitatively higher than anticipated based on review of individual case records and clinical experience. Specifically, while 97% sensitivity was assumed for colorimetric CO_2_ detection in the sample size calculation, both tests appeared to have a sensitivity of closer to 100%. This observation suggested that power to compare the tests may be lower than anticipated due to fewer false negatives from both tests and a smaller difference between the tests if both have near 100% sensitivity. Investigators did not have access to calculated sensitivity estimates or the difference in sensitivity between the two index tests. Accordingly, with respect to the pre-specified interim analysis above, the DSMB also evaluated (i) rates of missing index-test measurements (i.e., unpaired index test data), (ii) rates of indeterminate and non-tracheal reference standard determinations, and was asked to advise on whether the sample size should be re-estimated. After discussion with the DSMB, the target effect size for the primary comparison was revised to a 1-percentage-point absolute difference in sensitivity, representing the smallest difference likely to influence clinical practice or guideline recommendations (section 10 of the e-Appendix). Although a 1-percentage-point difference is small in relative terms when both tests approach perfect sensitivity, the absolute clinical effect of this difference is substantial at the population level. Given that approximately 1.5 million emergency intubations are performed annually in the United States^33,34^, a 1-percentage-point difference in sensitivity corresponds to roughly 15,000 additional intubations each year in which the less sensitive index test, if used, would yield a false negative result—possibly influencing a clinician to extubate a critically ill patient after the endotracheal tube had been correctly placed in the trachea and exposing that patient to unnecessary risks such as hypoxemia, aspiration, or airway injury. The required number of analyzable patients with a tracheal reference standard and paired index tests was recalculated to achieve ≥90% power to detect a 1–percentage point absolute difference in sensitivity at a two-sided α of 0.049.

To account for conditional dependence between the paired test results, the sample size re-estimation was conducted using the *power pairedprop* command in Stata (StataCorp, College Station, TX), assuming 99% sensitivity for waveform capnography, 98% sensitivity for colorimetric CO_2_ detection, two-sided α = 0.049, 90% power, and a conditional correlation between test results of 0.4, based on prior literature suggesting moderate dependence between paired diagnostic tests.^35^ Although the correlation between waveform capnography and colorimetric CO□ detection may be higher than 0.4, we assumed a slightly lower correlation to provide a more conservative sample size estimate, as lower correlations require a higher sample size. Under these assumptions, 1,946 patients with a tracheal reference standard and paired index test interpretations were required for the primary analysis.

To ensure adequate total enrollment, this analyzable sample size was inflated based on interim estimates of (a) the proportion of enrolled patients meeting the tracheal reference standard and (b) the proportion with paired index test interpretations. Based on observed rates of missing index test interpretations and indeterminate or non-tracheal reference standards at the interim analysis (approximately 7%), the revised target sample size was increased to 2,092 total enrolled patients.

While this sample size re-estimation was not pre-specified in the original protocol, simulation studies demonstrated that re-estimating the sample size using a smaller targeted effect size when the observed minimum sensitivity of the two index tests is greater than a given threshold would not be expected to inflate the type I error rate for the primary comparison of the two tests (section 10 of the e-Appendix). Similarly, these simulation studies also showed that re-estimating the sample size under these conditions would not bias the estimated sensitivity of the index tests. This modification was endorsed by the DSMB to ensure that the final sample size provides sufficient power to detect a difference that is small, yet still clinically meaningful, between two commonly used devices for determining endotracheal tube location during tracheal intubation of critically ill adults.

#### Additional Analyses of the Primary Outcome

*Sensitivity Analysis of the Primary Outcome: Accounting for Missing Index Test Interpretations* Instances when only one index test is performed and interpreted, rather than both, could reflect clinical circumstances that differ systematically from other patients (e.g., more urgent procedures). The main analysis (described above) will utilize a complete case analysis, which is consistent with missing completely at random. Because missingness may not be completely at random, we will also complete an analysis using multiple imputation using multiple chained equations implemented using the MICE package in R.

#### Sensitivity Analyses of the Primary Outcome: Accounting for Indeterminate Tube Location

For patients with a reference standard that is indeterminate as to whether the endotracheal tube is located in the trachea, we will conduct the following two sensitivity analyses:

1. All indeterminate assigned as tracheal: This represents the maximally inclusive scenario.
2. Clinician overall assessment: Indeterminate cases will be classified according to the clinician’s overall assessment of tube location recorded after the procedure. Cases judged to be in the trachea will be included in the sensitivity analysis; others will remain excluded.

For each reclassification scenario, we will analyze the data as described above. We note that the test statistic for McNemar’s test only uses counts of discordant pairs of index tests; thus, an indeterminate reference standard will only meaningfully affect the primary comparison when the results of the two index tests are discordant.

We will also present a table of narrative descriptions of cases with an indeterminate reference standard, including the corresponding index test interpretations, clinician assessment of the tube location, supporting evidence for this assessment, whether the tube was removed, whether cardiac arrest occurred, and additional pertinent clinical details.

*Analyses of heterogeneity of diagnostic test performance:*

Heterogeneity of diagnostic test performance refers to nonrandom variation in the accuracy of an index test across levels of a clinical characteristic. We will explore whether the sensitivity of waveform capnography and colorimetric CO_2_ detection differs across prespecified subgroups.

Because the overall sensitivity of both modalities is expected to be extremely high (∼98-99%) with sparse outcome variation, model-based interaction analyses (e.g., logistic regression with test-by-subgroup interaction terms) are unlikely to converge or yield stable estimates. Therefore, subgroup analyses will be descriptive and exploratory. Within each prespecified subgroup, we will directly estimate the sensitivity of each index test against the reference standard with exact 95% confidence intervals and calculate the absolute difference in sensitivity (with 95% confidence intervals). For continuous variables (e.g., systolic blood pressure), subgroups will be defined by quartiles.

The following variables available at the time of index test interpretation will be examined as potential effect modifiers: (1) location at enrollment (emergency department vs intensive care unit), (2) systolic blood pressure at the start of the intubation procedure (continuous), (3) active receipt of chest compressions at the time of index test interpretation (yes or no), (4) active problem of asthma or chronic obstructive pulmonary disease (yes or no), (5) vasopressor administration in the 2 min before or during the intubation procedure, (6) the presence of body fluids in the endotracheal tube after intubation, and (7) which index test was interpreted first. Finally, we will summarize each clinician’s cumulative prior experience interpreting waveform capnography and colorimetric CO_2_ detection, as reported at the time of enrollment, and display clinician experience for each modality with differentiation of concordant and discordant readings.

Additional details regarding heterogeneity of test performance are available in section 11 of e-Appendix 1.

#### Prespecified Analysis of Test Interpretation Order (Assessment of Non-Blinding)

Because complete blinding of the two clinicians responsible for index test interpretation was not possible, we will directly assess whether exposure to the other clinician’s interpretation influenced index-test performance. The order in which the two interpretations were solicited from clinicians was randomized 1:1, producing two randomized groups that differ only in which test was interpreted first. We will use this randomization to estimate the effect of interpretation order (i.e., the potential for the second clinician’s index test interpretation to be contaminated by knowledge of the first clinician’s index test interpretation) in two complementary ways.

First, and as the primary assessment of contamination, we will compare each index test’s sensitivity when it was interpreted first with its sensitivity when it was interpreted second. Because these groups comprise different patients, this is an unpaired comparison: for each test we will report the sensitivity (with exact 95% confidence intervals) in each order group and the absolute difference between the two, with 95% confidence intervals for the difference in independent proportions. A difference near zero indicates that knowledge of the other interpretation did not detectably alter that index test’s performance. These analyses are descriptive and exploratory.

Second, as described in the previous section, we report the absolute difference in sensitivity between the two index tests stratified by interpretation order.

#### Analysis of the Secondary Outcome

The main analysis of the secondary outcome will compare the specificity of waveform capnography with that of colorimetric CO_2_ detection among patients in whom both index tests were interpreted and the reference standard indicates a non-tracheal tube location. Because non-tracheal cases are expected to be rare, the analysis will focus on estimation rather than hypothesis testing. Specificity will be calculated using paired data only. Among reference-negative patients with paired index test interpretations, we will report the specificity of each test, the absolute difference in specificity, and the 95% confidence interval for the difference. If no discordant pairs are observed and both tests are 100% specific among reference-negative patients, the difference in specificity will be reported as 0, a paired confidence interval will be omitted as uninformative, and this circumstance will be explicitly noted.

Patients for whom the reference standard is indeterminate will not be categorized as indicating an endotracheal tube outside the trachea in the main analysis of the secondary outcome. To assess the robustness of this approach, we will perform similar sensitivity analyses as those used for the primary outcome; however, indeterminate cases will be considered as indicating an endotracheal tube that is outside the trachea so that they enter the denominator for specificity.

## Discussion

This manuscript describes the rationale, design, and prespecified statistical analysis plan for the WAVE study, a multicenter, pragmatic diagnostic accuracy study comparing waveform capnography and colorimetric CO detection for the determination of endotracheal tube location among critically ill adults undergoing tracheal intubation in the ED or ICU. By finalizing and publicly posting this plan prior to completion of enrollment and unblinded analysis, the WAVE study emphasizes transparency and analytic rigor.

Immediate confirmation that an endotracheal tube is located in the trachea is a critical step in emergency airway management. Failure to recognize non-tracheal (e.g., esophageal) endotracheal tube location can rapidly result in severe hypoxemia, cardiac arrest, and death. Conversely, unnecessary removal of a tube that is correctly located in the trachea—prompted by a false negative index test result—can expose critically ill patients to complications from repeated intubation attempts including hypoxemia, aspiration, and hemodynamic collapse. Both scenarios represent rare but potentially catastrophic failures in airway care.^1^

Waveform capnography and colorimetric CO_2_ detection are the two most used bedside tests for determining tube location in contemporary practice. Some existing guidelines recommend sole use of waveform capnography. However, many ED and ICU clinicians exclusively rely on colorimetric CO_2_ detection in their practice, highlighting the persistent uncertainty about comparative performance under the physiologic extremes encountered during a critical illness or injury.^10^ The WAVE study was designed to directly address this uncertainty by applying both tests to the same patients under real-world conditions and comparing their performance against a structured, non-CO_2_-based reference standard.

The sensitivity of each index test to accurately determine when the endotracheal tube is located in the trachea represents a genuine knowledge gap, particularly across the diverse physiologic conditions encountered in ED and ICU practice. Specificity, the probability that a test correctly identifies non-tracheal placement when the tube is located outside the trachea, remains a fundamental characteristic of any test determining endotracheal tube location. High specificity is critical because failure to recognize an esophageal intubation is life-threatening.

However, specificity was not selected as the primary outcome for several reasons. First, false-positive results, when a test indicates tracheal placement despite esophageal location, are extraordinarily rare for both waveform capnography and colorimetric CO_2_ detection, occurring only under unusual circumstances described in case reports.^19,36,37^ Second, given the low contemporary prevalence of esophageal intubation, meaningful differences in specificity between the two tests are unlikely to be measurable in a pragmatic study of feasible size. The inability to determine differences in specificity reflects the success of modern airway practice rather than a deficiency of the study design. Importantly, treating specificity as a secondary outcome does not diminish its clinical importance. Instead, this analytic structure acknowledges that when non-tracheal intubation occurs, both tests are expected to perform nearly perfectly, and the primary remaining source of uncertainty—and potential patient harm—lies in false-negative interpretations when the tube is actually in the trachea. The WAVE study therefore prioritizes sensitivity while still rigorously estimating specificity and transparently reporting cases in which discordant or unexpected results occur.

This study has important limitations. The most notable is that complete blinding is not possible. The lack of blinding applies to both the reference standard and index tests, each of which has different risks of bias.

The first form, assessing the reference standard with knowledge of the index-test results, is empirically associated with overestimated accuracy in single-test studies with subjective reference standards, but poses minimal risk in WAVE. The reference standard relies on objective endpoints (exhaled tidal volume, bronchoscopic visualization, or 20 min of sustained spontaneous circulation without tube removal) that cannot be swayed by observer awareness of the index tests. Furthermore, even if diagnostic review bias were possible, meta-epidemiologic data demonstrate that its effect is modest (median relative diagnostic odds ratio ∼1.1-1.3)^38,39^, which at the expected sensitivities of 97–99% corresponds to well under one percentage point of absolute inflation for each test, and should not meaningfully affect the comparison in sensitivities between the two tests.

The second form, a clinician interpreting an index test who may be exposed to the other device or to the other clinician’s verbalized interpretation, is a residual limitation we could not fully eliminate despite all the measures described in the *Blinding* subsection. However, several considerations indicate that its effect on the primary comparison is negligible. Under the QUADAS-C framework for comparative accuracy studies, a limitation shared by both index tests biases the comparison between them only insofar as it acts differentially between the two tests.^40^ Because the order of interpretation was randomized 1:1, exposure between devices is structurally balanced; symmetric contamination pulls the tests toward mutual agreement, acting as a conservative bias toward the null that attenuates rather than creates a difference. While asymmetric contamination could theoretically both widen or attenuate the observed difference depending on which discordant readings are influenced, only attenuation toward the null threatens the study’s ability to detect a true difference. Formal tipping-point modeling indicates that for a true sensitivity difference of ≥1.5 percentage points, attenuation below the clinically important 1 percentage-point threshold requires a contamination rate of 33% or higher under symmetric contamination, and 21.7% or higher under the theoretical worst case scenario where contamination selectively turns negative colorimetric readings to positive (details in Section 12 of the e-Appendix). Reducing a true difference to zero under this worst-case scenario would require extreme contamination (≥65%). At a plausible contamination fraction of 10%, only narrow true differences (1.00 to 1.11 percentage points symmetrically, or 1.00 to 1.20 asymmetrically) could be attenuated below the clinically important threshold; larger true differences remain above it, and reclassification within this narrow band is itself clinically inconsequential. Finally, because the order in which the test interpretations were solicited was randomized 1:1, we can assess the effect of any contamination empirically. In a prespecified analysis, we will compare each index test’s sensitivity when it was interpreted first with its sensitivity when it was interpreted second; a difference near zero provides direct evidence that exposure to the other interpretation did not meaningfully affect performance.

Differential expectation bias could theoretically arise if the intubating clinician directly observed tube passage between the vocal cords while a non-intubating interpreter did not. However, this asymmetry is largely neutralized by the routine use of video laryngoscopy across participating units, which projects the airway view onto an external display and affords both bedside clinicians shared visualization of tube passage. Furthermore, while index-test assignment was determined before the procedure, the order in which interpretations were solicited was randomized 1:1, preventing either modality from systematically anchoring the assessment. Preserving access to these routine procedural observations mirrors real-world airway management and adheres to QUADAS guidance that diagnostic tests should be evaluated within their intended clinical context.^31^

An additional limitation is that CO_2_-based diagnostic tests for determining endotracheal tube position during emergency intubation cannot necessarily distinguish a tube whose tip is located below the vocal cords within the thoracic trachea (correct position) from a tube whose tip is located at or just above the vocal cords (incorrect position, but not esophageal). In both scenarios, with the endotracheal tube cuff inflated, waveform capnography or the colorimetric CO_2_ detector may capture alveolar ventilation and sustained exhaled CO_2_ from the endotracheal tube. However, these index tests may falsely reassure the clinician that the airway has been secured when, in fact, the tube tip is at or above the vocal cords. Furthermore, because the reference standard operationalizes tracheal placement primarily through effective pulmonary ventilation, its ventilation criteria may also be satisfied if delivered tidal volume is directed through a malpositioned tube perched at or just above the vocal cords.^19^ An endotracheal tube perched at or just above the vocal cords is dangerous because there is a high likelihood of further dislodgement and ventilatory failure in the short term; fortunately, this scenario is rare. To the extent that such cases exist, both index tests and the reference standard are likely to indicate that the tube is located in the trachea. The primary comparison of sensitivity between the two index tests is therefore unlikely to be biased by this phenomenon, though absolute sensitivity estimates for both tests may be marginally inflated.

## Interpretation

In summary, the WAVE study was deliberately designed to address a clinically meaningful and unresolved question in emergency airway management: how waveform capnography and colorimetric CO_2_ detection compare in their ability to reliably determine endotracheal tube location among critically ill adults, many with deranged cardiorespiratory physiology. By prioritizing sensitivity as the primary outcome, rigorously estimating specificity as a secondary outcome, and applying a robust, non-CO_2_-based reference standard, this prespecified analysis plan aligns methodological rigor with real-world clinical decision-making. The results of the WAVE study will provide clinicians, guideline authors, and health systems with contemporary evidence to inform best practice in the selection of exhaled CO_2_ detection devices for high-risk airway management.

## Data Availability

All data produced in the present study will be available upon reasonable request to the authors

## References

1. Chrimes N, Higgs A, Hagberg CA, et al. Preventing unrecognised oesophageal intubation: a consensus guideline from the Project for Universal Management of Airways and international airway societies. Anaesthesia. 2022;77(12):1395–1415.

2. Hansel J, Law JA, Chrimes N, Higgs A, Cook TM. Clinical tests for confirming tracheal intubation or excluding oesophageal intubation: a diagnostic test accuracy systematic review and meta-analysis. Anaesthesia. 2023;78(8):1020–1030.

3. Ward KR, Yealy DM. End-tidal carbon dioxide monitoring in emergency medicine, Part 1: Basic principles. Acad Emerg Med. 1998;5(6):628–636.

4. Ornato JP, Shipley JB, Racht EM, et al. Multicenter study of a portable, hand-size, colorimetric end-tidal carbon dioxide detection device. Ann Emerg Med. 1992;21(5):518–523.

5. Rabitsch W, Nikolic A, Schellongowski P, et al. Evaluation of an end-tidal portable ETCO2 colorimetric breath indicator (COLIBRI). Am J Emerg Med. 2004;22(1):4–9.

6. Jaffe MB. Respiratory gas analysis-technical aspects. Anesth Analg. 2018;126(3):839–845.

7. Falk JL, Rackow EC, Weil MH. End-Tidal Carbon Dioxide Concentration during Cardiopulmonary Resuscitation. N Engl J Med. 1988;318(10):607–611.

8. Thompson JE, Jaffe MB. Capnographic waveforms in the mechanically ventilated patient. Respir Care. 2005;50(1):100–108; discussion 108-109.

9. Li J. Capnography alone is imperfect for endotracheal tube placement confirmation during emergency intubation. J Emerg Med. 2001;20(3):223–229.

10. Bunting AJ, Driver BE, Ilg AM, et al. Use of waveform capnography and colorimetric CO detection for tracheal intubation: A National Survey of ED and ICU practices. Am J Emerg Med. 2026;104:190–192.

11. Prekker ME, Driver BE, Trent SA, et al. Video versus direct laryngoscopy for tracheal intubation of critically ill adults. N Engl J Med. 2023;389(5):418–429.

12. Russotto V, Myatra SN, Laffey JG, et al. Intubation Practices and Adverse Peri-intubation Events in Critically Ill Patients From 29 Countries. JAMA. 2021;325(12):1164–1172.

13. Law JA, Duggan LV, Asselin M, et al. Canadian Airway Focus Group updated consensus-based recommendations for management of the difficult airway: part 1. Difficult airway management encountered in an unconscious patient. Can J Anaesth. 2021;68(9):1373–1404.

14. Wigginton JG, Agarwal S, Bartos JA, et al. Part 9: Adult advanced life support: 2025 American Heart Association guidelines for cardiopulmonary resuscitation and emergency cardiovascular care. Circulation. 2025;152(16_suppl_2):S538–S577.

15. Myatra SN, Shah AP, Ramkumar V, et al. All India Difficult Airway Association 2025 Guidelines for the management of unanticipated difficult airway in adults under general anaesthesia. Indian J Anaesth. 2025;69(11):1117–1141.

16. Ahmad I, El-Boghdadly K, Iliff H, et al. Difficult Airway Society 2025 guidelines for management of unanticipated difficult tracheal intubation in adults. Br J Anaesth. Published online November 7, 2025. doi:10.1016/j.bja.2025.10.006

17. Apfelbaum JL, Hagberg CA, Connis RT, et al. 2022 American Society of Anesthesiologists Practice Guidelines for Management of the Difficult Airway. Anesthesiology. 2022;136(1):31–81.

18. Nellcor Adult Colorimetric CO2 Detector, Directions for Use. Accessed March 20, 2026. https://www.manualslib.com/manual/1674525/Covidien-Nellcor-Easycap-Ii.html

19. Driver B, Law J. Confirmation of correct endotracheal tube placement in adults. UpToDate. Accessed June 3, 2026. https://www.uptodate.com/contents/confirmation-of-correct-endotracheal-tube-placement-in-adults

20. Sum-Ping S, Mehta M, Anderton J. A comparative study of methods of detection of esophageal intubation. Anesth Analg. 1989;69:627–632.

21. León MA, Räsänen J, Mangar D. Neural Network-Based Detection of Esophageal Intubation. Anesthesia & Analgesia. 1994;78(3):548.

22. Raphael DT, Crookes P, Arnaudov D, Benbassat M. Acoustic reflectometry esophageal profiles minimally affected by massive gas ventilation. Am J Emerg Med. 2005;23(6):747–753.

23. Bair AE, Laurin EG, Schmitt BJ. An assessment of a tracheal tube introducer as an endotracheal tube placement confirmation device. Am J Emerg Med. 2005;23(6):754–758.

24. Farmery AD, Roe PG. A model to describe the rate of oxyhaemoglobin desaturation during apnoea. Br J Anaesth. 1996;76(2):284–291.

25. Mort TC. The incidence and risk factors for cardiac arrest during emergency tracheal intubation: a justification for incorporating the ASA Guidelines in the remote location. J Clin Anesth. 2004;16(7):508–516.

26. Stein P, Albrecht R, Spahn DR. Multiple trauma, resuscitation, and 15 minutes of esophageal intubation: survival without neurologic deficit. American Journal of Emergency Medicine. 2014;32(8):950–e1.

27. Timmermann A, Russo SG, Eich C, et al. The out-of-hospital esophageal and endobronchial intubations performed by emergency physicians. Anesth Analg. 2007;104(3):619–623.

28. Honardar MR, Posner KL, Domino KB. Delayed Detection of Esophageal Intubation in Anesthesia Malpractice Claims: Brief Report of a Case Series. Anesth Analg. 2017;125(6):1948–1951.

29. Simel DL, Rennie D, Bossuyt PMM. The STARD statement for reporting diagnostic accuracy studies: application to the history and physical examination. J Gen Intern Med. 2008;23(6):768–774.

30. Bossuyt PM, Leeflang MM. Cochrane Handbook for Systematic Reviews of Diagnostic Test Accuracy Version 0. 2008.

31. Davenport CF, Rutjes AWS, Mallett S, et al. QUADAS-3 explanation and elaboration: Guidance for quality assessment of diagnostic test accuracy studies. Ann Intern Med. 2026;179(4):e2504943.

32. Trajman A, Luiz RR. McNemar chi2 test revisited: comparing sensitivity and specificity of diagnostic examinations. Scand J Clin Lab Invest. 2008;68(1):77–80.

33. Pfuntner A, Wier LM, Stocks C. Most frequent procedures performed in US hospitals, 2011: statistical brief# 165. In: Healthcare Cost and Utilization Project (HCUP) Statistical Briefs. (http://www.ncbi.nlm.nih.gov/books/NBK174682/). Agency for Healthcare Research and Quality; 2013.

34. Cairns C, Kang K. National Hospital Ambulatory Medical Care Survey: 2021 emergency department summary tables. https://www.cdc.gov/nchs/data/nhamcs/web_tables/2021-nhamcs-ed-web-tables-508.pdf

35. Leisenring W, Alonzo T, Pepe MS. Comparisons of predictive values of binary medical diagnostic tests for paired designs. Biometrics. 2000;56(2):345–351.

36. Asai T, Shingu K. Case report: A normal capnogram despite esophageal intubation. Can J Anaesthesia-journal Can D Anesthesie. 2001;48:1025–1028.

37. MacLeod BA, Heller MB, Gerard J, Yealy DM, Menegazzi JJ. Verification of endotracheal tube placement with colorimetric end-tidal CO2 detection. Ann Emerg Med. 1991;20(3):267–270.

38. Lijmer JG, Mol BW, Heisterkamp S, et al. Empirical evidence of design-related bias in studies of diagnostic tests. JAMA. 1999;282(11):1061–1066.

39. Rutjes AWS, Reitsma JB, Di Nisio M, Smidt N, van Rijn JC, Bossuyt PMM. Evidence of bias and variation in diagnostic accuracy studies. CMAJ. 2006;174(4):469–476.

40. Yang B, Mallett S, Takwoingi Y, et al. QUADAS-C: A tool for assessing risk of bias in comparative Diagnostic Accuracy Studies. Ann Intern Med. 2021;174(11):1592–1599.

