## Supplementary Appendix for "Protocol and Statistical Analysis Plan for the Waveform Capnography versus Colorimetric Carbon Dioxide Detection During Tracheal Intubation of Critically Ill Adults (WAVE) Study"


**1. List of WAVE Investigators** 1
**2. STARD 2015 Checklist** 3
Table S1: STARD 2015 Checklist 3
**3. Baseline Index Test Use, By Study Site** 7
**SUPPLEMENTARY METHODS** 8
**4. Ethics and Informed Consent Processes** 8
Risks of Tracheal Intubation in the ED or ICU 8
Potential Risks of Participation in the WAVE Study 8
Potential Benefits of Participation in the WAVE Study 9
Minimization of Risk 10
Waiver of Consent 10
Provision of Information after Participation 12
**5. Data collection forms** 14
**6. Reference standard determination** 17
**7. Rationale for the Choice of the Primary Outcome** 18
**8. Rationale for the Choice of the Secondary Outcome** 20
**9. Composition and Responsibilities of the Data Safety Monitoring Board** 21
**10. Details of Sample Size Calculation** 22
**11. Analyses of Heterogeneity of diagnostic test performance** 28
**12. Contamination tipping-point analysis** 30
**13. Safety Monitoring and Adverse Events** 34
**14. Plan for Communication of Protocol Changes** 35
**15. Patient Privacy, Data Storage, and Sharing** 36
**16. Study Status** 37
**REFERENCES** 38

### 1. List of WAVE Investigators

BLINDED

### 2. STARD 2015 Checklist

Table S1: STARD 2015 Checklist

| **Section / Item** | **Item No** | **Description** | **Addressed on Page Number** |
|---|---|---|---|
| **Title / Abstract** | 1 | Identification as a diagnostic accuracy study (e.g., sensitivity, specificity, AUC) | 1 |
| | 2 | Structured summary of study design, methods, results, and conclusions | 1 |
| **Introduction** | 3 | Scientific and clinical background, intended use and role of the index test | 3 |
| | 4 | Study objectives and hypotheses | 5-6 |
| **Methods – Study Design & Participants** | 5 | Whether data collection was prospective or retrospective | 6 |
| | 6 | Eligibility criteria | 7 |
| | 7 | Basis for identifying participants (e.g., symptoms, screening test) | 7 |
| | 8 | Setting, location, and dates | 7 |
| | 9 | Participant recruitment (consecutive, random, convenience) | 7 |
| **Methods – Test Methods** | 10a | Index test(s) described in sufficient detail to allow replication | 9-11 |

| | | | |
|---|---|---|---|
| | 10b | Reference standard described in sufficient detail to allow replication | 11-14 |
| | 11 | Rationale for choosing reference standard | 11-14 |
| | 12a | Definition & rationale for index test cutoffs/thresholds | 9-11 |
| | 12b | Definition & rationale for reference standard cutoffs/thresholds | 11-14 |
| | 13 | Whether test performers/readers were blinded to other results | 14-15 |
| **Methods – Analysis** | 14 | Methods for estimating diagnostic accuracy | 18-19 |
| | 15 | Handling of indeterminate results | 22-23 |
| | 16 | Handling of missing data | 22-23 |
| | 17 | Methods for evaluating variability in diagnostic accuracy (subgroups, sensitivity analyses) | 23-24 |
| | 18 | Whether analyses were pre-specified or exploratory | 18 |
| **Results – Participants** | 19 | Flow diagram of participants | To be presented in final results manuscript |

| | | | |
|---|---|---|---|
| | 20 | Baseline characteristics of participants | To be presented in final results manuscript |
| | 21 | Distribution of disease severity & alternative diagnoses | To be presented in final results manuscript |
| | 22 | Whether sample reflects clinical population | To be presented in final results manuscript |
| **Results – Test Results** | 23 | Cross-tabulation of index test vs reference standard | To be presented in final results manuscript |
| | 24 | Diagnostic accuracy estimates (sens, spec, AUC, CI) | To be presented in final results manuscript |
| | 25 | Adverse events | To be presented in final results manuscript |
| **Discussion** | 26 | Limitations, potential bias, generalizability | 25-29 |
| | 27 | Implications for practice | 25-29 |
| **Other Information** | 28 | Registration number and registry name | 7 |

| | | | |
|---|---|---|---|
| | 29 | Where full protocol can be accessed | To be presented in final results manuscript |
| | 30 | Funding sources and role of funders | 6 |

### 3. Baseline Index Test Use, By Study Site

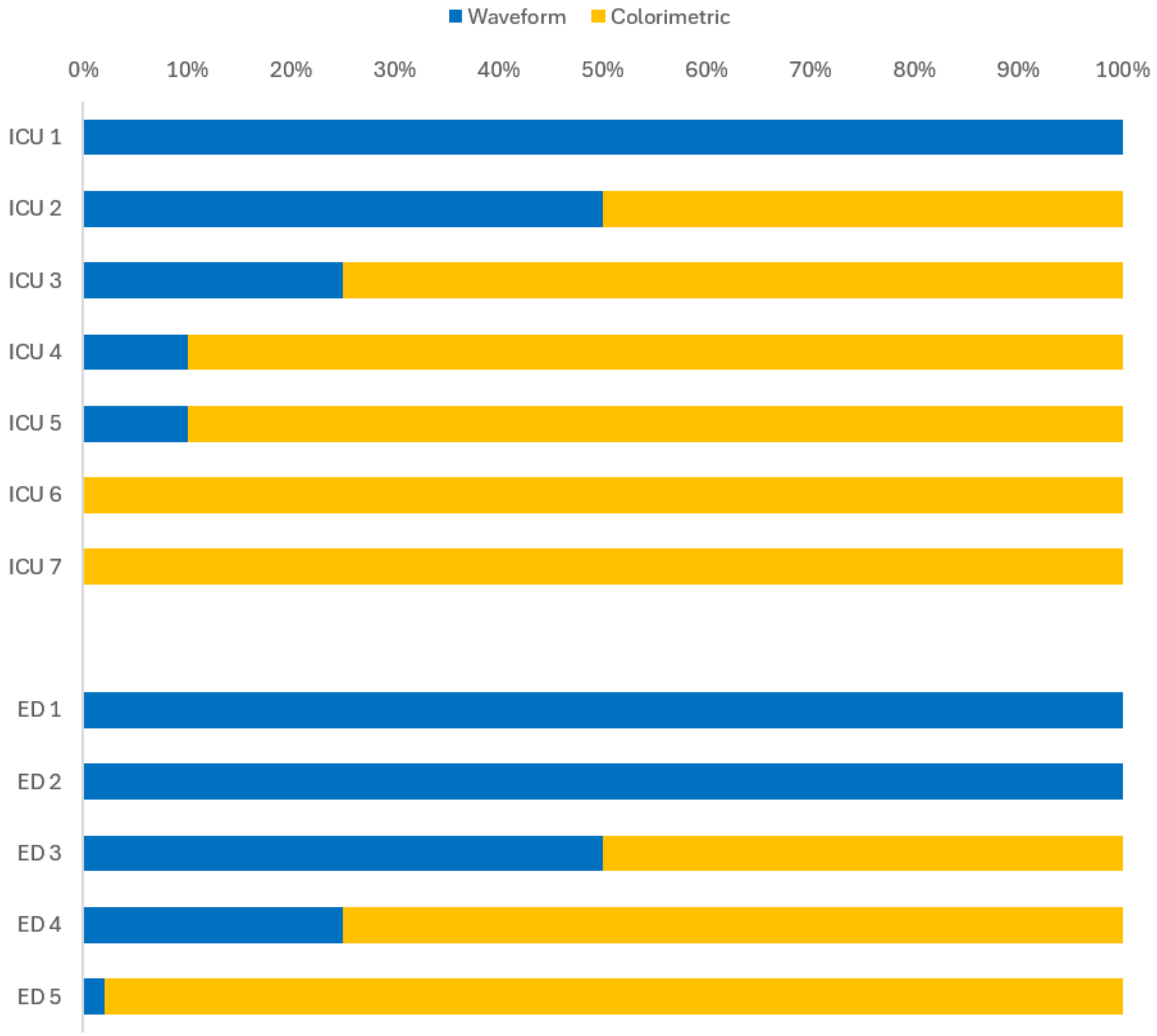


**Figure S1. Percentage use of waveform capnography and colorimetric CO2 detection at baseline (before the start of the WAVE study) at the 7 ICUs and 5 EDs.** Three sites used exclusively waveform capnography (blue), 2 sites used exclusively colorimetric $CO_2$ detection (orange), and the remaining 7 sites used a mixture of both index tests.

### SUPPLEMENTARY METHODS

#### 4. Ethics and Informed Consent Processes

Risks of Tracheal Intubation in the ED or ICU

Patients who are severely ill enough to require emergency tracheal intubation in the ED or ICU as part of their clinical care are at high risk of complications related to the procedure. Many patients are undergoing intubation for hypoxemia or hemodynamic instability. Severe hypoxemia or cardiovascular instability occurs during nearly half of intubations in the ED and ICU.[1–3] Hypoxemia and hypotension during intubation are associated with an increased risk of cardiac arrest and death.[4] Cardiac arrest occurs in approximately 1-in-25 cases of emergency tracheal intubation in adults.[3] Other complications during intubation may include aspiration (approximately 2.8% of cases), esophageal intubation (1.3%), injury to oral or dental structures (0.2%), and pneumothorax (0.1%). The long-term consequences of complications occurring during emergency tracheal intubation are unclear. Neurologic recovery from traumatic brain injury may be worse after peri-procedural hypoxemia due to a secondary ischemic insult.[5]

Potential Risks of Participation in the WAVE Study

Participation in the WAVE study involves minimal incremental risk because:

- All patients eligible for the study are already undergoing tracheal intubation with a plan for determination of endotracheal tube location using an exhaled $CO_2$ test as part of their clinical care.
- Waveform capnography and colorimetric $CO_2$ detection are both standard-of-care interventions that are in widespread clinical use.[6]
- No benefits or risks are currently known to substantially differ between the two tests when used to determine the location of the endotracheal tube during tracheal intubation.

- Both tests have been determined to be acceptable options for the optimal care of the patient by treating clinicians (otherwise the patient is excluded from the study).
- The final location of the index endotracheal tube will be assessed with both exhaled $CO_2$ tests rather than just one test, decreasing the chance of a false negative or false positive single test triggering an unnecessary intervention.
- When the two tests result in discrepant interpretations by the clinicians (i.e., one test is consistent with the tube in the trachea but the other test is not consistent with a tube in the trachea), the clinician is alerted to the possibility of an endotracheal tube that is not located in the trachea. The clinician will do additional evaluation, potentially at an earlier time point than if a sole exhaled $CO_2$ test had registered a false positive or false negative result. A discrepancy between waveform capnography and colorimetric $CO_2$ detection for a tube located in the trachea will not typically result in unnecessary tube removal in stable patients. Rather, the usual practice if the clinician believes that the trachea was intubated (i.e., they visualized the tube passing between the vocal cords) is to seek additional clinical evidence of correct tube location using repeat laryngoscopy, thoracic ultrasound, or bronchoscopy before removing the endotracheal tube and starting over.

Although no risks are currently known to differ between waveform capnography and colorimetric $CO_2$ detection, it is possible that the results of the WAVE study will ultimately demonstrate a difference between the two approaches in the accuracy of determining the location of the endotracheal tube. However, patients are receiving both tests during the study and any difference between the two is unlikely to affect patient care.

Potential Benefits of Participation in the WAVE Study

The primary benefits of the WAVE study will be the indirect benefits to society that would result if one test to determine the location of the endotracheal tube is found to be superior to the other.

Because millions of critically ill adults undergo emergency tracheal intubation each year, if one of the two tests were found to have superior sensitivity, implementation of that finding in clinical practice would potentially improve the care provided to millions of severely ill patients.

Compared to the minimal risks of participation in the study, the pursuit of these benefits is reasonable.

Minimization of Risk

Federal regulations 45 CFR 46.111(a)(1) require that risks to patients are minimized by using procedures which are consistent with sound research design. This trial meets this human subjects protection requirement by incorporating numerous design elements to minimize risk to patients.

Both tests to determine the location of the endotracheal tube have been used in clinical practice for decades in the same populations eligible to participate in the WAVE study. To further mitigate risk, we exclude patients for whom treating clinicians determine that one $CO_2$-based test is required or contraindicated for the optimal care of the patient.

Finally, to limit the risks associated with the collection of protected health information (PHI), the minimum amount of PHI necessary for study conduct will be collected. The data will be coded and stored in a secure online database (REDCap) only accessible by the investigators. REDCap tools will be used to ensure that the PHI that is collected is only visible to investigators at the healthcare system where the patient is enrolled. To protect participant privacy, REDCap tools will be used to ensure that only deidentified data can be exported for analysis by the investigators.

Waiver of Consent

Waveform capnography and colorimetric $CO_2$ detection are both standard-of-care tests to determine the location of the endotracheal tube during tracheal intubation.[7,8] Because of the paucity of definitive data showing the superiority of one test over the other, there is variation in the use of these tests. This study will enroll patients who are undergoing emergency tracheal intubation as part of their clinical care for whom treating clinicians have determined that they will determine the tube location with exhaled $CO_2$ testing. Each enrolled patient will receive both tests simultaneously (paired diagnostic test accuracy design).

This study has been approved with a waiver of informed consent because the study involves minimal incremental risk and obtaining informed consent would be impracticable.

Obtaining informed consent is impracticable because:

- The expected medical condition of patients requiring emergency tracheal intubation in the ED or ICU is critical. Based on prior trials in the same patient population and setting, approximately 70% of patients eligible for this trial will be experiencing acute encephalopathy (altered mental status) due to their critical illness. The anticipated median Glasgow coma scale score is 11 (equivalent to moderate brain injury). Among the minority of patients whose level of consciousness is not impaired, 45-55% will be experiencing acute delirium. Thus, most patients eligible for the trial will not have the capacity to provide informed consent. Further, family members or legally authorized representatives (LARs) are frequently unavailable during the short time interval between the decision to perform tracheal intubation and the start of the procedure in critically ill patients in the ED or ICU.
- The time available for patients or LARs to consider participation will be insufficient. Even when a patient retains decisional capacity or a LAR is immediately available, a meaningful informed consent process is precluded by the rapid clinical events leading up to emergency tracheal intubation. No published literature has quantified the time from clinicians' decision to perform emergency tracheal intubation (the inclusion criteria for this trial) until the completion of the intubation procedure (when the $CO_2$-based index

tests are interpreted). Historically, more than 50% of intubations occurred within 5 minutes after treating clinicians verbalized the decision to intubate or ordered an induction medication. Obtaining informed consent for research requires study personnel to assess decisional capacity, identify a LAR when appropriate, review the informed consent document in a quiet setting, and provide sufficient time for the patient or LAR to process the information, assess the risks and benefits of participation, and ask questions. Meaningful informed consent cannot be executed in the 5 minutes between the decision to perform emergency tracheal intubation and initiation of the procedure. Emergency tracheal intubation of critically ill adults is a time-sensitive procedure for which every minute of delay increases the likelihood of hypoxemia, hypotension, and periprocedural cardiac arrest. Delaying emergency tracheal intubation to attempt a meaningful informed consent process would be unsafe, impracticable, and unethical.

Because this observational cohort study involves minimal incremental risk, the study would not adversely affect the welfare or privacy rights of the participant, and obtaining informed consent would be impracticable, the IRBs at participating sites have approved this protocol and waived the requirement for informed consent. This consent approach is consistent with the conduct of multiple prior randomized clinical trials that included critically ill patients undergoing emergency tracheal intubation.[8–13]

Provision of Information after Participation

45 CFR 46.116(f)(3)(v) states that “Whenever appropriate, the subjects or legally authorized representatives will be provided with additional pertinent information after participation”. Information regarding the study will be made available to patients and families by one of three mechanisms: (1) a patient and family notification sheet provided to each patient and family following enrollment informing the patient of his or her enrollment and describing the study, (2) a

patient and family information sheet posted in at least three publicly-visible locations within the study unit containing general information about the study and contact information for the research team for additional questions or concerns, (3) a patient and family information sheet provided to each patient and family on admission as part of an “admission packet” containing general study information and contact information for the research team for additional questions or concerns. Which mechanism of providing information to patients and families will be used at each study site will be determined by site investigators in coordination with the local context assessment of the site IRB.

### 5. Data collection forms

***Page 1 of 2***

1st Version of page 1: STEP 2 is colorimetric detector interpretation.

Patient label here

**STEP 1:** **Vital signs at start of 1st intubation attempt**

1. $O_2$ sat as meds pushed: ________ % or ☐ $O_2$ sat not available
2. SBP as meds pushed: ________ mmHg or ☐ SBP not available
3. HR as meds pushed: ________ bpm or ☐ HR not available
4. End-tidal $CO_2$ as meds pushed: ________ mmHg or ☐ $EtCO_2$ not available

**STEP 2: COLORIMETRIC DETECTOR** after 7 breaths

**Ask clinician #1:** *Is the color change consistent with a tube located in the trachea?*

☐ Yes ☐ No ➞ If No, what was observed? ______________________________

-Should change from purple to yellow (expiration) and back to purple (inspiration)
-If color is tan during expiration, give 6 more breaths and mark "yes" if remains tan or changes to yellow

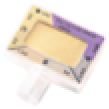

**STEP 3: WAVEFORM CAPNOGRAPHY** after 7 breaths

**Ask clinician #2:** *Is the waveform consistent with a tube located in the trachea?*

☐ Yes ☐ No ➞ If No, what was observed? ______________________________

-Waveform rises during expiration and falls during inspiration
-Amplitude is consistent or increasing over 7 breaths
-Peak amplitude is >7.5 mmHg above baseline

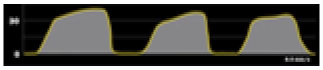

**STEP 4:** **AFTER INTUBATION**

1. End-tidal CO2 value ________ mmHg after 7 breaths or ☐ $EtCO_2$ value not available
2. Was the patient receiving CPR when $CO_2$ was interpreted (during STEP 2 and STEP 3)?
   ☐ Yes ☐ No
3. 1st SBP after procedure ________ mmHg or ☐ SBP value not available

**STEP 5:** **Confirm with clinician:**

**1. Mechanical ventilation effective through original tube?**
For 5 consecutive ventilations, measured exhaled volume ≥ 80% of delivered tidal volume
→ ☐ **Yes** → STOP Turn page over
↓ **No**

**2. Original tube removed within 20 min?** → ☐ **Yes** →
↓ **No**

**3. Death or ongoing CPR at 20 min?** → ☐ **Yes** →
↓ **No**

Where do you believe original tube was located?
☐ In trachea ☐ Not in trachea
Why do you believe tube located there? (select all that apply)
☐ Bronchoscopy
☐ Video laryngoscopy
☐ Thoracic or tracheal ultrasound
☐ Esophageal detector device (Wee)
☐ Other ________________
→ STOP Turn page over

**4. Patient alive AND original tube is not removed at 20 min?** → ☐ **Yes** → STOP Turn page over

2nd version of page 1: STEP 2 is waveform capnography interpretation.

**STEP 1: Vital signs at start of 1st intubation attempt**

1. $O_2$ sat as meds pushed: ________ % or ☐ $O_2$ sat not available
2. SBP as meds pushed: ________ mmHg or ☐ SBP not available
3. HR as meds pushed: ________ bpm or ☐ HR not available
4. End-tidal $CO_2$ as meds pushed: ________ mmHg or ☐ $EtCO_2$ not available

Patient label here

**STEP 2: WAVEFORM CAPNOGRAPHY** after 7 breaths

**Ask clinician #1:** *Is the waveform consistent with a tube located in the trachea?*

☐ Yes ☐ No ➞ If No, what was observed? ______________________________

-Waveform rises during expiration and falls during inspiration
-Amplitude is consistent or increasing over 7 breaths
-Peak amplitude is >7.5 mmHg above baseline

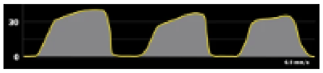

**STEP 3: COLORIMETRIC DETECTOR** after 7 breaths

**Ask clinician #2:** *Is the color change consistent with a tube located in the trachea?*

☐ Yes ☐ No ➞ If No, what was observed? ______________________________

-Should change from purple to yellow (expiration) and back to purple (inspiration)
-If color is tan during expiration, give 6 more breaths and mark "yes" if remains tan or changes to yellow

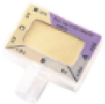

**STEP 4: AFTER INTUBATION**

1. End-tidal CO2 value ________ mmHg after 7 breaths or ☐ $EtCO_2$ value not available
2. Was the patient receiving CPR when $CO_2$ was interpreted (during STEP 2 and STEP 3)?
   ☐ Yes ☐ No
3. 1st SBP after procedure ________ mmHg or ☐ SBP value not available

**STEP 5: Confirm with clinician:**

**1. Mechanical ventilation effective through original tube?**
For 5 consecutive ventilations, measured exhaled volume ≥ 80% of delivered tidal volume
☐ Yes → STOP Turn page over
No ↓

**2. Original tube removed within 20 min?**
☐ Yes → (see box below) → STOP Turn page over
No ↓

**3. Death or ongoing CPR at 20 min?**
☐ Yes → (see box below) → STOP Turn page over
No ↓

**Where do you believe original tube was located?**
☐ In trachea ☐ Not in trachea
**Why do you believe tube located there?** (select all that apply)
☐ Bronchoscopy
☐ Video laryngoscopy
☐ Thoracic or tracheal ultrasound
☐ Esophageal detector device (Wee)
☐ Other ______________________

**4. Patient alive AND original tube is not removed at 20 min?**
☐ Yes → STOP Turn page over

**Clinician completes after intubation**

1. What was the primary indication for intubation?
   ☐ **Inability to protect airway** ☐ **Failure to oxygenate** ☐ **Cardiac arrest**
   ☐ **Failure to ventilate** ☐ **Procedure** ☐ **Hemodynamic instability**

2. Was patient receiving CPR at the time of first laryngoscopy attempt? ☐ **Yes** | ☐ **No**

3. Active medical problems at the time of intubation (select all that apply):
   ☐ **Cardiac arrest without ROSC** ☐ **Asthma** ☐ **COPD** ☐ **Systolic CHF**
   ☐ **Cardiac arrest with ROSC** ☐ **Pulmonary embolism** ☐ **DKA or acidosis** ☐ **Shock**

4. Select all the locations that body fluids were present at the time of intubation and tube confirmation:
   ☐ **Mouth** | ☐ **Endotracheal tube** | ☐ **Colorimetry device** | ☐ **Waveform device** | ☐ **No body fluids were present**

5. Did the patient receive a vasopressor in the 2 min before the $CO_2$ assessment? ☐ **Yes** | ☐ **No**

6. Did you have to troubleshoot the waveform capnography? ☐ **Yes** | ☐ **No**

6a. If yes, what did you do? ☐ **Plug in tubing** | ☐ **Unplug and replug tubing** | ☐ **Replace sidestream adapter**
(select all that apply) ☐ **Replace monitoring module** | ☐ **Change monitor settings** | ☐ **Other:**____________

6b. If yes, did troubleshooting work to restore the $CO_2$ waveform? ☐ **Yes** | ☐ **No**

6c. If yes, approx. time spent troubleshooting $CO_2$ waveform? ☐ **<30 sec** | ☐ **30-59 sec** | ☐ **1-2 min** | ☐ **>2 min**

7. Did you have to troubleshoot the colorimetry device? ☐ **Yes** | ☐ **No**

7a. If yes, what did you do? ☐ **Replace color device** | ☐ **Clean device** | ☐ **Other:**__________________________

7b. If yes, did troubleshooting work to get the colorimetry device to work? ☐ **Yes** | ☐ **No**

7c. If yes, approx. time spent troubleshooting colorimetry? ☐ **<30 sec** | ☐ **30-59 sec** | ☐ **1-2 min** | ☐ **>2 min**

8. If the tube was removed within 20 minutes, why was it removed? ☐ **N/A, tube not removed**
   ☐ **Esophageal or other non-tracheal placement** | ☐ **ETT cuff or pilot balloon not holding air** | ☐ **Other ETT malfunction**
   ☐ **Patient self-extubation** | ☐ **Inadvertent extubation** | ☐ **Other:** ________________________

*Suppose* you couldn't confirm this tube with exhaled $CO_2$ and you removed it after intubation:

9. Would you have been able to rapidly and reliably ventilate the lungs with a facemask? ☐ **Yes** | ☐ **No**

10. Would you have been able to rapidly and reliably ventilate the lungs with a supraglottic airway? ☐ **Yes** | ☐ **No**

11. Would the patient have been at high risk of aspirating a substantial amount of body fluids into the lungs? ☐ **Yes** | ☐ **No**

**Clinician #1:** **First and Last Name:** ______________________________________

Specialty (circle one): **Emergency Medicine / Critical Care / Anesthesia / Other:** __________________

Training level (circle one): **Attending / Fellow / Resident / CRNA / NP / PA / RT / Other:** ___________________

Estimated # of times you have used waveform capnography to confirm intubation previously: _________

Estimated # of times you have used colorimetric $CO_2$ detector to confirm intubation previously:________

**Clinician #2:** **First and Last Name:**_______________________________________

Specialty (circle one): **Emergency Medicine / Critical Care / Anesthesia / Other:** __________________

Training level (circle one): **Attending / Fellow / Resident / CRNA / NP / PA / RT / Other:** __________________

Estimated # of times you have used waveform capnography to confirm intubation previously: __________

Estimated # of times you have used colorimetric $CO_2$ detector to confirm intubation previously: ________

### 6. Reference standard determination

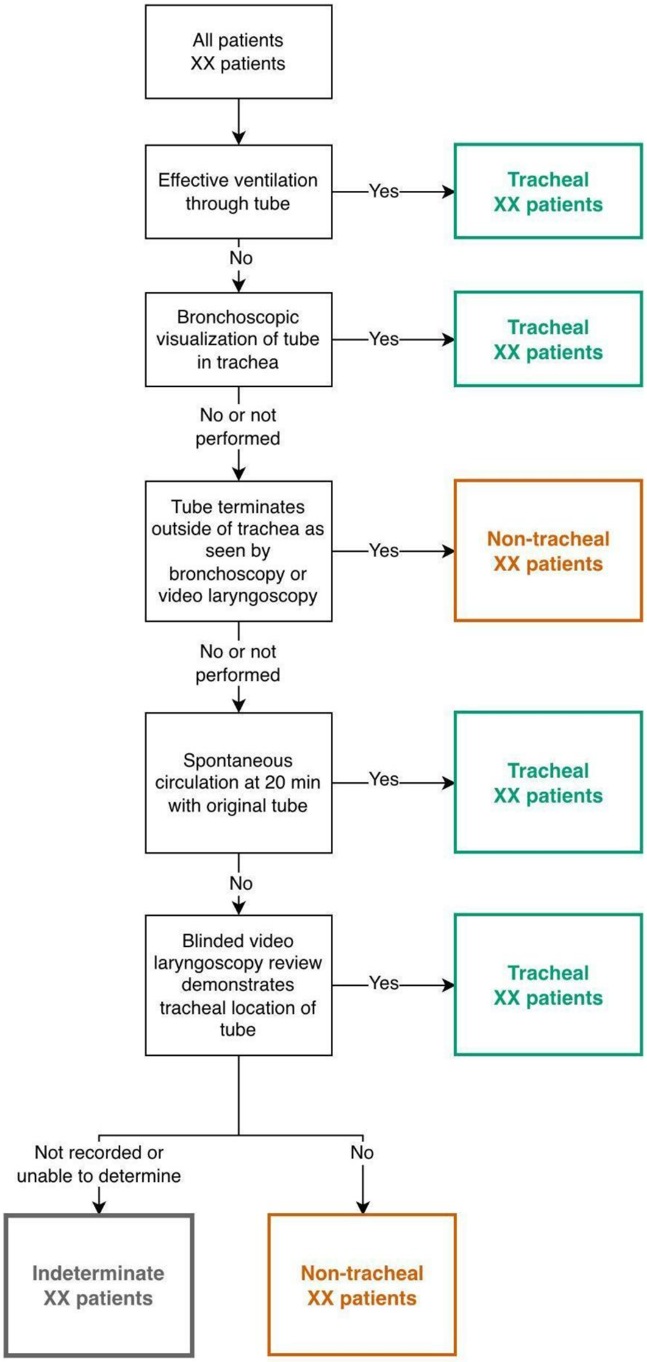


**Figure S2:** This flow diagram shows how patients are assigned a reference standard for the location of the endotracheal tube. Patients who meet neither a tracheal nor non-tracheal reference standard are assigned a value of “indeterminate”.

### 7. Rationale for the Choice of the Primary Outcome

The primary outcome is the sensitivity of the index tests (waveform capnography and colorimetric $CO_2$ detection) for determining when the endotracheal tube is located in the trachea. Sensitivity, in this context, refers to the proportion of endotracheal tubes located in the trachea in which the index test interpretation is consistent with a tube located in the trachea (i.e., the true positive rate). Sensitivity was chosen as the primary endpoint for two principal reasons.

First, sensitivity is clinically meaningful and has advantages over specificity as a primary outcome. The specificity of both index tests to identify a tube *outside of the trachea* is universally accepted to be essentially perfect.[14–25] False-positive results, which are instances in which the index test indicates tracheal placement for a tube located outside of the trachea, occur only under rare and unusual circumstances. These include when, (1) the distal tube is located at or just above the vocal cords, (2) there is an abnormal conduit between the trachea and the esophagus (e.g., tracheoesophageal fistula), or (3) when a spontaneously breathing patient has a tube in the esophagus with an uninflated cuff.[26,27] Because the index tests perform almost perfectly except in these rare conditions, meaningful differences in specificity between waveform and colorimetric detection are unlikely to be measurable in a pragmatic observational study of this size, assuming an esophageal intubation rate of ≤ 1% (as observed in prior trials from this network[7,28]).

Second, the sensitivity of these index tests in contemporary emergency and critical care practice is an important knowledge gap. Although data are limited, clinicians have speculated that pathophysiologic alterations in pulmonary blood flow or alveolar ventilation, such as cardiac arrest, profound shock, or severe bronchospasm, might lower the sensitivity of one or both index tests. The American Heart Association notes that "In patients with decreased pulmonary blood flow from low cardiac output or cardiac arrest, $ETCO_2$ may not be as reliable."[29]

Some clinicians believe exhaled $CO_2$ detection is frequently unreliable based on these abnormal physiologic states frequently encountered in critically ill adults. The clinical consequences of this belief are illustrated in inquests into deaths and malpractice reviews involving unrecognized esophageal intubation: in these cases, clinicians attributed an absent exhaled $CO_2$ signal to patient physiology (eg, bronchospasm, pneumothorax, hypovolemia, severe pneumonia, pericarditis, or cardiac arrest) rather than to tube misplacement, misinterpreting a correct test result as a false negative caused by reduced sensitivity.[30–32] This pattern suggests that uncertainty about sensitivity in pathophysiologic states leads clinicians to discount a negative $CO_2$ result, potentially delaying recognition of esophageal intubation with fatal consequences. Existing evidence on this point is limited and based largely on exhaled $CO_2$ tests that are no longer available and populations studied three decades ago. Available data suggest that while the sensitivity of both waveform capnography and colorimetric $CO_2$ detection is, in general, near 100%[14–17,19,21,27], some studies report modestly lower sensitivities (~98%) for both tests in non-cardiac arrest scenarios,[18,20] and lower sensitivities in the setting of cardiac arrest, especially for colorimetric devices.[18–20,22,27,33] Only one study has reported 100% sensitivity for waveform capnography in the setting of cardiac arrest.[17]

No contemporary study has systematically examined waveform capnography and colorimetric $CO_2$ detection in ED and ICU patients, including many patients with bronchospasm, acidemia, traumatic injury, hypotension, and shock. If one test has a lower sensitivity, it would be interpreted as not consistent with an endotracheal tube in the trachea more often when the tube is actually in the trachea. This would potentially lead to immediate and unnecessary tube removals for fear of esophageal intubation, especially if the patient is hypoxemic or unstable. This scenario would expose patients to unnecessary risk relative to use of the other index test, including hypoxemia, aspiration, and complications from repeated intubation attempts.

Accordingly, the WAVE study was designed to determine the sensitivities of both waveform capnography and colorimetric $CO_2$ detection across diverse clinical scenarios and real-world conditions, in part to identify subgroups in which sensitivity may be reduced.

**8. Rationale for the Choice of the Secondary Outcome**

The secondary outcome is the specificity of the index tests (waveform capnography and colorimetric $CO_2$ detection) for identifying endotracheal tubes located outside the trachea. Specificity represents the proportion of endotracheal tubes terminating outside the trachea for which the index test correctly indicates a non-tracheal location via the absence of sustained exhaled $CO_2$ (the true negative rate). Accurate detection of non-tracheal placement is a fundamental purpose of exhaled $CO_2$ testing because prompt recognition and removal of an esophageal intubation is lifesaving.

Although specificity is the more clinically consequential characteristic because undetected esophageal intubation is immediately life-threatening, it was not selected as the primary outcome because the near-perfect specificity of both tests is already established and unlikely to differ meaningfully between devices in a study of this size.[14–25] Sensitivity, by contrast, represents a genuine knowledge gap, particularly across the diverse physiologic conditions encountered in ED and ICU practice. Specificity will be assessed as a secondary outcome to confirm expected performance in a contemporary population.

**9. Composition and Responsibilities of the Data Safety Monitoring Board**

The Data and Safety Monitoring Board (DSMB) consists of members with expertise in emergency medicine, pulmonary medicine, critical care medicine, biostatistics, and clinical research. All members of the DSMB are voting members. The DSMB developed a charter and approved the trial monitoring plan prior to the interim analysis. The DSMB has the ability at any point to recommend that the study end, be modified, or continue unchanged.

The principal role of the DSMB is to assure the safety of patients in the study. The DSMB monitors data, reviews and assesses the performance of its operations, and makes recommendations to the Steering Committee with respect to:

- Review of adverse events
- Interim results of the study
- Possible early termination of the study because of new external information, early attainment of study objectives, safety concerns, or inadequate performance
- Possible modifications in the study protocol
- Performance of individual centers

### 10. Details of Sample Size Calculation

Initial Sample Size Estimate

The initial version of the sample size estimate, included in the study protocol submitted to the institutional review board on January 16, 2025, is described here.

The sensitivity of waveform capnography to verify tracheal tube location is approximately 99% according to published literature.[15–17,34–36] For colorimetric $CO_2$ detection, sensitivity is also high and has been reported to be between 97-100%.[19,27,37] A survey of clinicians at the sites within the research network performing the trial indicated that an absolute difference in sensitivity of 1-2 percentage points between waveform capnography and colorimetric $CO_2$ detection would be clinically meaningful. A difference of 2% in a procedure as common as emergency tracheal intubation would represent an unnecessary tube removal or additional testing in 2 out of every 100 intubations. In the United States, there are at least 1.5 million emergency tracheal intubations performed annually.[38,39]

Assuming a sensitivity of 99% for waveform capnography, a sample size of 1,294 patients would be needed to determine whether there is at least a 2% absolute difference in sensitivity between the two index tests with 95% statistical power and a two-sided alpha level of 0.05. Accounting for incomplete data or non-tracheal tube location in up to 3% of patients, we planned to enroll 1,332 patients. These calculations were performed using the Stata command *power pairedprop* and assumed independence of the two index test results among patients with tracheal tube placement. Because these calculations did not explicitly account for conditional correlation between the paired diagnostic tests, they were likely substantially conservative.

Final Sample Size Estimate

The final version of the sample size estimate, included in the study protocol submitted on March 13, 2026, after the pre-planned interim analysis of the WAVE study, is described here.

After discussion with the independent Data and Safety Monitoring Board (DSMB) at the pre-specified interim analysis, the original sample size was re-estimated to ensure adequate power for the final primary analysis. First, the required number of analyzable patients with a tracheal reference standard and paired index tests was recalculated to achieve ≥90% power to detect a 1–percentage point absolute difference in sensitivity at two-sided $\alpha$=0.049. A 1-percentage-point difference in sensitivity was the smallest difference that participating clinicians indicated would alter practice patterns or recommendations, and was felt to be clinically meaningful because a difference of ≥1% would represent at least 15,000 intubation procedures per year, out of 1.5 million that occur outside of the operating room[39,40], for which the endotracheal tube location would not be correctly determined by the index test. Second, this analyzable sample size was inflated to total enrollment based on observed interim estimates of (a) the proportion of enrolled patients meeting the tracheal reference standard and (b) the proportion with paired, interpretable index-test results.

The sample size calculation used the *power pairedprop* command in Stata (StataCorp, College Station, TX) with assumptions of 99% sensitivity for waveform capnography, 98% sensitivity for colorimetric $CO_2$ detection, two-sided $\alpha$ = 0.049, 90% power, and conditional correlation between test results of 0.4 (based on prior literature in which paired diagnostic technologies often exhibit moderate dependence). Although the correlation between waveform capnography and colorimetric $CO_2$ detection may be higher than 0.4, we assumed a slightly lower correlation to provide a more conservative sample size estimate, as lower correlations require a higher sample size.

Under these assumptions, 1,946 patients with a tracheal reference standard and paired index test results are required for the primary analysis of sensitivity. Based on observed rates of missing paired index test interpretations and indeterminate or non-tracheal reference standards at the interim analysis (~7%), the DSMB recommended targeting 2,092 total enrolled patients to ensure that the required analyzable sample is reached.

This modification was endorsed by the DSMB to ensure that the final sample size provides sufficient power to detect a difference that is small, yet still clinically meaningful, between two commonly used modalities for determining endotracheal tube location.

No calculation for power was made for the secondary outcome, specificity, as a reference standard of a non-tracheal tube location will be rare and both index tests are expected to have specificity approaching 100% (e-Appendix sections 7 and 8).

Simulations

The following simulations assess whether the unplanned sample size re-estimation inflated the type I error rate or biased sensitivity estimates. In short, it did not. As described in the main document, the DSMB was asked to consider whether the sample size of the study should be re-estimated after study investigators noticed that the sensitivities of both index tests were likely closer to 100% than originally anticipated. In particular, the sensitivity of colorimetric $CO_2$ detection was originally assumed to be 97%, implying that power to compare the tests may be lower than planned, due to fewer false negatives from each test and a smaller difference between tests (as neither can have sensitivity greater than 100%). While this re-estimation was not pre-specified, we conducted simulations evaluating the potential impact on the type I error rate. Specifically, while study investigators did not have access to calculated sensitivity rates for either test, the decision to re-estimate the sample size was operationally similar to if they had chosen to do so based on the *minimum sensitivity of the two tests being greater than or equal to a given threshold—in this case, approximately 98%.*

We first evaluated the potential impact of the sample size re-estimation on the type I error rate for the primary analysis by simulating data under the null hypothesis of there being no difference in the sensitivity of the two index tests. In all cases, the original sample size of 1,294 (prior to inflation to account for loss-to-follow up) was used as the initial sample size; this was then re-estimated using updated sensitivities of 99% (waveform capnography) and 98%

(colorimetric $CO_2$ detection), *only if the minimum of the observed sensitivities of the two index tests was ≥ 98%*. The updated sensitivities used in the sample size re-estimation were determined in practice based on the minimum difference that would be clinically important, not the observed rates, which the study investigators did not have access to; these therefore remained fixed across simulations. These simulations used a significance-level of 0.05 as they did not include an interim analysis of efficacy and 100,000 replications were used in all cases. The type I error rate was estimated by the proportion of the 100,000 replications in which McNemar's test rejected the null hypothesis of no difference in the sensitivity of the two tests. The results for type I error when varying the true sensitivities of both index tests from 95% to 99% are shown in Figure S3. For all values of the sensitivity of each index test, the type I error rate remains at the specified 5% both marginally and conditional on re-estimating the sample size.

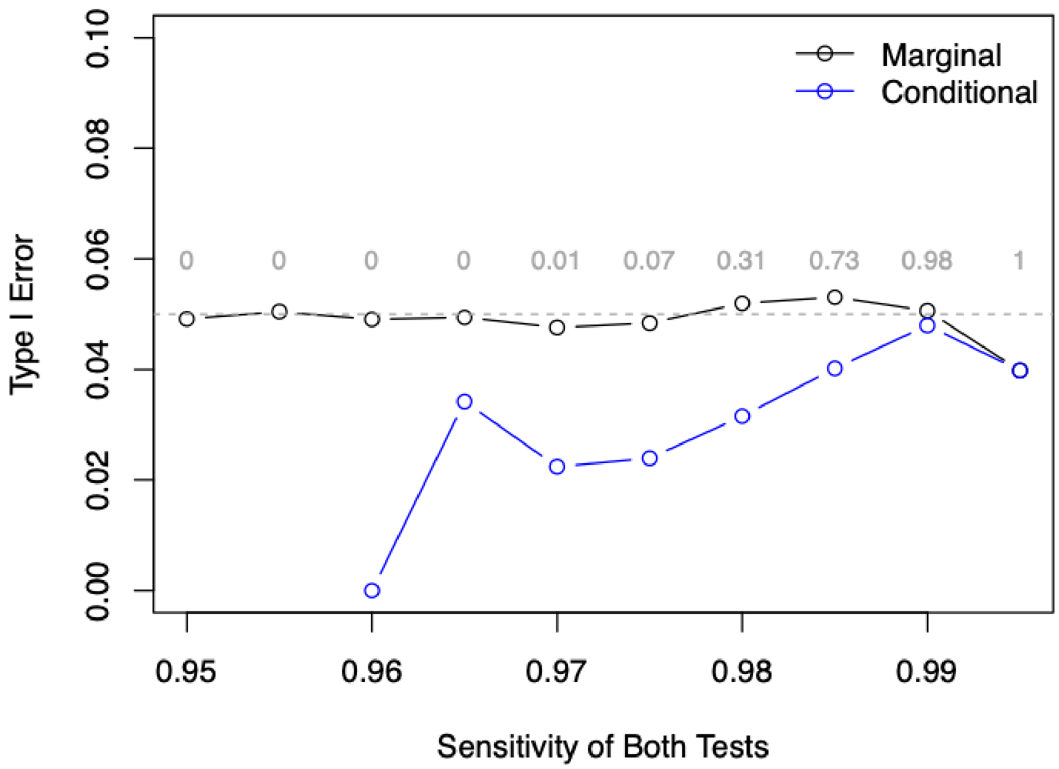

**Figure S3.** Simulated type I error rate for McNemar's test across a range of sensitivity of both index tests. Black dashed line shows type I error rate marginally, over all simulation replications. Blue dashed line shows type I error rate conditional on re-estimating the sample size (minimum observed sensitivity at 50% enrollment ≥ 98%). The gray numbers depict the proportion of simulation iterations in which the sample size was re-estimated.

Table S2 shows the mean estimated sensitivity for each test across the 100,000 simulation replications. This demonstrates that under the null hypothesis, the sample size re-estimation procedure is not expected to bias the estimated sensitivity of either index test.

| **True Sensitivity of Both Index Tests** | **0.95** | **0.955** | **0.96** | **0.965** | **0.97** | **0.975** | **0.98** | **0.985** | **0.99** | **0.995** |
|---|---|---|---|---|---|---|---|---|---|---|
| Mean Estimated Sensitivity of Waveform Capnography | 0.950 | 0.955 | 0.960 | 0.965 | 0.970 | 0.975 | 0.980 | 0.985 | 0.990 | 0.995 |
| Mean Estimated Sensitivity of Colorimetric $CO_2$ Detection | 0.950 | 0.955 | 0.960 | 0.965 | 0.970 | 0.975 | 0.980 | 0.985 | 0.990 | 0.995 |
| Re-estimation Rate | 0.00 | 0.00 | 0.00 | 0.00 | 0.01 | 0.07 | 0.31 | 0.73 | 0.98 | 1.00 |
| Re-estimation Count | 0 | 0 | 12 | 117 | 982 | 7,355 | 30,558 | 72,811 | 97,518 | 99,997 |

**Table S2.** Mean estimated sensitivity for each index test, as well as rate and count of sample size re-estimation, across 100,000 simulation replications under the null hypothesis for a range of true sensitivities of both index tests.

We next simulated data under the alternative hypothesis where the sensitivities of the two index tests differ (Table S3). In all scenarios, the sensitivity of waveform capnography was fixed at 99% and the sensitivity of colorimetric $CO_2$ detection was varied from 95% to 99.5%. The table below demonstrates that the sample size re-estimation procedure is also not expected to bias the estimated sensitivity of either index test under any of these scenarios.

| True Sensitivity of Colorimetric $CO_2$ Detection | 0.95 | 0.955 | 0.96 | 0.965 | 0.97 | 0.975 | 0.98 | 0.985 | 0.99 | 0.995 |
|---|---|---|---|---|---|---|---|---|---|---|
| Mean Estimated Sensitivity of Waveform Capnography | 0.990 | 0.990 | 0.990 | 0.990 | 0.990 | 0.990 | 0.990 | 0.990 | 0.990 | 0.990 |
| Mean Estimated Sensitivity of Colorimetric CO2 Detection | 0.950 | 0.955 | 0.960 | 0.965 | 0.970 | 0.975 | 0.980 | 0.985 | 0.990 | 0.995 |
| Re-estimation Rate | 0.00 | 0.00 | 0.00 | 0.01 | 0.05 | 0.18 | 0.46 | 0.81 | 0.97 | 0.98 |
| Re-estimation Count | 0 | 21 | 172 | 1,030 | 4,741 | 17,804 | 46,479 | 81,467 | 97,123 | 98,474 |

**Table S3.** Mean estimated sensitivity for each index test, as well as rate and count of sample size re-estimation, across 100,000 simulation replications under a range of true sensitivity of colorimetric $CO_2$ detection. The sensitivity of waveform capnography is fixed at 99% in all scenarios.

**11. Analyses of Heterogeneity of diagnostic test performance**

In accordance with the Instrument for assessing the Credibility of effect Modification Analyses (ICEMAN) recommendations,[41] we have prespecified the following variables available at the time of index test interpretation as potential effect modifiers and hypothesized the direction of effect modification for each:

a. Location of intubation (ED vs ICU): We hypothesize that location of intubation will not have an effect on the primary outcome.
b. Systolic blood pressure at time of intubation (analyzed in quartiles): If dichotomized for presentation in a forest plot then we will present systolic blood pressure as greater than or equal to 100 mm Hg versus less than 100 mm Hg. We hypothesize that a lower systolic blood pressure will decrease the sensitivity of the colorimetric $CO_2$ detector relative to the sensitivity of waveform capnography. This is based on the physiologic rationale that arterial hypotension can be associated with decreased pulmonary blood flow, especially in cardiogenic or hypovolemic shock. Less pulmonary blood flow would deliver less $CO_2$ to the alveolar-capillary membrane, potentially decreasing alveolar $CO_2$ partial pressure below the threshold of detection for the colorimetric $CO_2$ detector (the threshold for yellow color change is 15 mm Hg) but not the waveform capnograph.
c. Receipt of chest compressions during index test interpretation (Yes vs No): We hypothesize that in the setting of active chest compressions during index test interpretation, the sensitivity of colorimetric $CO_2$ detection will be reduced as compared with waveform capnography. This is based on physiologic rationale that reduction in pulmonary blood flow and $CO_2$ production during cardiac arrest, even accounting for high-quality cardiopulmonary resuscitation, may result in an attenuated but still detectable amplitude of waveform capnography whereas the end-tidal $CO_2$ concentration may not reach the threshold for detection by colorimetric $CO_2$ detector (~15 mm Hg).

d. Active problem of asthma or chronic obstructive pulmonary disease (Yes vs No): We hypothesize that bronchospasm (airflow obstruction due to asthma or chronic obstructive pulmonary disease) will not have an effect on the primary outcome, as pulmonary blood flow is not typically affected and $CO_2$ should be detected if any ventilation is occurring.
e. Vasopressor administration during the 2 min before and during tracheal intubation (Yes vs No): We hypothesize that vasopressor administration will not have an effect on the primary outcome.
f. The presence of body fluids within the lumen of the endotracheal tube after intubation (Yes vs No): We hypothesize that the presence of body fluids within the endotracheal tube will decrease the sensitivity of waveform capnography more than it will for the colorimetric detector because the tubing connecting the device to the monitor can be obstructed by fluid, impairing accurate transmission of exhaled gas to the sensor.
g. Order of test interpretation (colorimetric $CO_2$ detection first vs waveform capnography first): We hypothesize that the order of test interpretation will not have an effect on the primary outcome.

Clinician prior experience: We hypothesize that the clinician's cumulative prior experience with the index test will not have an effect on the primary outcome. This analysis is separate from the preceding analyses and will not be included in a forest plot. We will use patient-level data to graphically display clinician experience for the test that that clinician interpreted, differentiating concordant and discordant instances.

### 12. Contamination tipping-point analysis

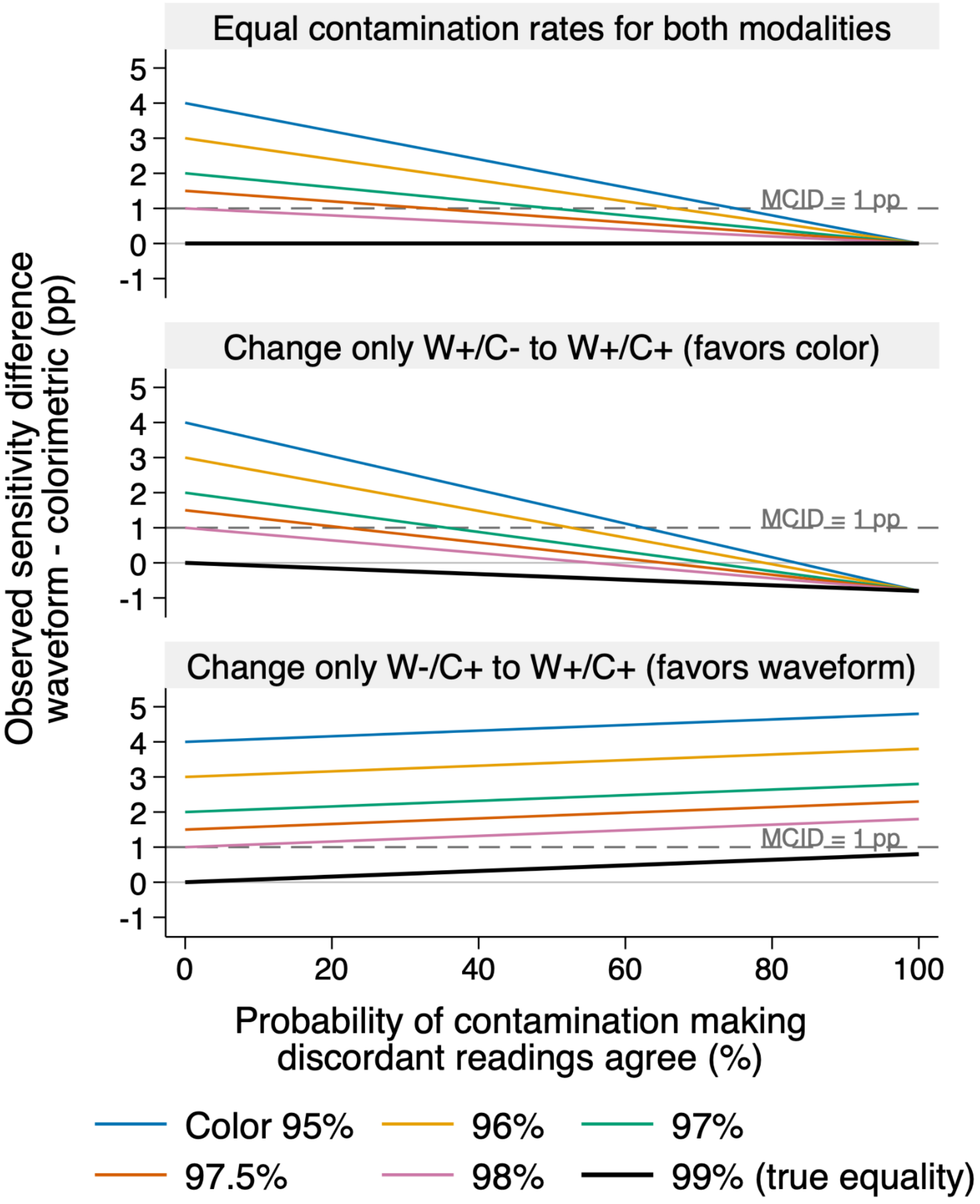


**Figure S4. Contamination tipping-point analysis.**

In Fig. S4, each line shows the expected observed difference in sensitivity between waveform capnography and colorimetric $CO_2$ detection (waveform minus colorimetric) under specified contamination assumptions. True (uncontaminated) waveform sensitivity is assumed to be 99%, and true colorimetric sensitivities range from 95% to 99%. The probability that both tests would yield negative interpretations without contamination is assumed to be 0.2% among patients with a reference standard of tube located in the trachea. The dashed gray line marks the minimal clinically important difference of 1 percentage point (pp); the thin solid gray line marks zero difference.

The horizontal axis represents contamination, the probability of one test adopting the other's reading whenever the two would otherwise disagree, turning a discordance into a concordance. Contamination was modeled as resolving discordance toward concordance for tracheal placement (both waveform and colorimetric detection positive for tracheal placement), reflecting the clinical reality that operators encountering one positive test may actively scrutinize the second device for confirmation of successful intubation. Converting a positive reading to negative based on an adjacent negative test is much less likely to occur.

Let *b* and *c* denote the uncontaminated probabilities of waveform-positive/colorimetric-negative and waveform-negative/colorimetric-positive pairs, respectively. If the probabilities that contamination turns a discordant pair into a concordant one are defined as $g_b$ and $g_c$, the expected observed sensitivity difference is $(1-g_b)b - (1-g_c)c$. Letting $\gamma$ denote the contamination probability plotted on the horizontal axis, the three panels set $(g_b, g_c)$ to $(\gamma,\gamma)$, $(\gamma,0)$, and $(0,\gamma)$, respectively. Equal-rate contamination attenuates the observed difference by a factor of $1-\gamma$.

The top panel assumes equal probabilities of resolving waveform-positive/colorimetric-negative and waveform-negative/colorimetric-positive discordances.. The middle panel demonstrates the scenario in which only waveform-positive/colorimetric-negative pairs are made concordant by contamination, which increases colorimetric detection sensitivity without affecting waveform sensitivity. The bottom panel demonstrates the scenario in which only waveform-negative/colorimetric-positive pairs are made concordant by contamination, increasing waveform capnography sensitivity without affecting colorimetric detection sensitivity. When discordant values become concordant for one modality more than the other, this can attenuate or exaggerate the observed difference in sensitivity between the two tests. For other contamination scenarios not shown above, expected sensitivity differences are bounded by the two selective-resolution scenarios shown in the bottom two panels.

Under the equal-rate contamination scenario (top panel), the observed difference in sensitivity reaches the 1-percentage-point threshold at contamination probabilities of 75%, 67%, 50%, and 33% for true (uncontaminated) colorimetric sensitivities of 95%, 96%, 97%, and 97.5%, respectively. At a more plausible contamination fraction of 10%, only a true difference between 1.00 and 1.11 percentage points could be attenuated below the threshold; larger true differences remain above it.

Under the scenario favoring colorimetric $CO_2$ detection (middle panel), where only discordant negative colorimetric readings turn positive, and thus concordant with waveform, the observed difference in sensitivity reaches the 1-percentage-point threshold at contamination probabilities of 62.5%, 52.6%, 35.7%, and 21.7% for true (uncontaminated) colorimetric sensitivities of 95%, 96%, 97%, and 97.5%, respectively.  A true sensitivity difference of exactly 1 percentage point, corresponding to colorimetric sensitivity of 98%, falls below the threshold with any contamination. At a more plausible contamination fraction of 10%, only a true difference between 1.00 and 1.20 percentage points could be attenuated to the threshold or below; larger true differences remain above it.

Under the scenario where contamination selectively converts discordant negative waveform readings to positive (bottom panel), the observed sensitivity difference favoring waveform capnography widens. This scenario cannot obscure a true sensitivity advantage for waveform capnography, although it could exaggerate its magnitude or create an apparent advantage when uncontaminated sensitivities are equal.

Thus, the worst-case scenario for attenuating a true difference to below the 1-percentage-point threshold occurs when contamination is completely one-sided: converting discordant negative colorimetric readings to positive. In this scenario, for an assumed true difference of 1.5 percentage points (colorimetric sensitivity 97.5%), attenuation below the threshold occurs only if the contamination probability is 21.7% or higher. We consider contamination of this magnitude unlikely given the instructions to view only the assigned device, commit to an interpretation, and because the order of solicitation was randomized. The randomized solicitation order provides an empirical assessment of order-related effects. Small estimated differences with confidence intervals excluding clinically important order effects would provide reassurance against this mechanism of contamination.

### 13. Safety Monitoring and Adverse Events

Assuring patient safety is an essential component of this protocol. Determining the location of the endotracheal tube with waveform capnography or colorimetric $CO_2$ detection devices falls within the standard of care and have been used in clinical practice for decades with an established safety profile. However, any research conducted during a high-risk, time-sensitive procedure like tracheal intubation of critically ill adults raises unique safety considerations. This protocol addresses these considerations through:

1. Systematic collection of outcomes relevant to the safety of determining the location of the endotracheal tube with both waveform capnography and colorimetric $CO_2$ detection.
2. Structured monitoring, assessment, recording, and reporting of adverse events.
3. Periodic review every 4 months of all instances of tube removal, cardiac arrest, or death within 20 minutes of intubation where one or both of the index tests was interpreted as consistent with non-tracheal tube location. The study safety monitor determines if it is plausible that the discordant interpretation of the index tests (i.e. one test interpreted as consistent with a tube located in the trachea and the other test interpreted as not consistent with a tube located in the trachea) was contributory to the tube removal, cardiac arrest, or death. This study safety monitor has the ability to recommend that the study end, be modified, or continued unchanged at each review interval.

**14. Plan for Communication of Protocol Changes**

Any changes to the study protocol, including changes to eligibility, outcomes, and analyses, will be implemented via a new version of the full study protocol, tracked with the date of the update, and the version number of the study protocol. A list summarizing the changes made with each protocol revision will be included at the end of each protocol. The updated protocol will be sent to the IRB for approval prior to implementation of the protocol change. At the time of publication, the original study protocol and the final study protocol, including the summary of changes made with each protocol version, will be included in the supplementary material for publication.

### 15. Patient Privacy, Data Storage, and Sharing

Federal regulations 45 CFR 46.111 (a) (7) requires that, when appropriate, there are adequate provisions to protect the privacy of patients and to maintain the confidentiality of data. At no time during the course of this study, its analysis, or its publication will patient identities be revealed in any manner. The minimum necessary data containing patient or provider identities will be collected. All patients will be assigned a unique study ID number for tracking. All data collected for this study will be entered into a secure online database. Tools within the secure online database will be used so that only the coordinating center and investigators from the enrolling site will have access to data from patients enrolled at that site. All data will be maintained in the secure online database until the time of study publication. At the time of publication, a de-identified version of the database will be generated. Deidentified data will be available for sharing following study publication with the requirements and terms of data sharing specified in the study results manuscript.

### 16. Study Status

The WAVE study is a pragmatic, prospective, paired diagnostic accuracy study comparing waveform capnography and colorimetric $CO_2$ detection to a reference standard. Enrollment began on April 21, 2025, and is expected to conclude in 2026.

### REFERENCES


1. Martin LD, Mhyre JM, Shanks AM, Tremper KK, Kheterpal S. 3,423 emergency tracheal intubations at a university hospital: airway outcomes and complications. *Anesthesiology*. 2011;114(1):42-48.

2. Jaber S, Jung B, Corne P, et al. An intervention to decrease complications related to endotracheal intubation in the intensive care unit: a prospective, multiple-center study. *Intensive Care Med*. 2010;36(2):248-255.

3. Russotto V, Myatra SN, Laffey JG, et al. Intubation Practices and Adverse Peri-intubation Events in Critically Ill Patients From 29 Countries. *JAMA*. 2021;325(12):1164-1172.

4. April MD, Arana A, Reynolds JC, et al. Peri-intubation cardiac arrest in the Emergency Department: A National Emergency Airway Registry (NEAR) study. *Resuscitation*. 2021;162:403-411.

5. Rubenson Wahlin R, Nelson DW, Bellander BM, Svensson M, Helmy A, Thelin EP. Prehospital Intubation and Outcome in Traumatic Brain Injury—Assessing Intervention Efficacy in a Modern Trauma Cohort. *Front Neurol*. 2018;9. doi:10.3389/fneur.2018.00194

6. Bunting AJ, Driver BE, Ilg AM, et al. Use of waveform capnography and colorimetric $CO_2$ detection for tracheal intubation: A National Survey of ED and ICU practices. *Am J Emerg Med*. 2026;104:190-192.

7. Prekker ME, Driver BE, Trent SA, et al. Video versus Direct Laryngoscopy for Tracheal Intubation of Critically Ill Adults. *N Engl J Med*. 2023;389(5):418-429.

8. Driver BE, Semler MW, Self WH, et al. Effect of Use of a Bougie vs Endotracheal Tube With Stylet on Successful Intubation on the First Attempt Among Critically Ill Patients Undergoing Tracheal Intubation: A Randomized Clinical Trial. *JAMA*. 2021;326(24):2488-2497.

9. Driver BE, Prekker ME, Moore JC, Schick AL, Reardon RF, Miner JR. Direct versus video laryngoscopy using the C-MAC for tracheal intubation in the emergency department, a randomized controlled trial. *Acad Emerg Med*. 2016;23(4):433-439.

10. Driver BE, Prekker ME, Klein LR, et al. Effect of use of a bougie vs endotracheal tube and stylet on first-attempt intubation success among patients with difficult airways undergoing emergency intubation: A randomized clinical trial. *JAMA*. 2018;319(21):2179-2189.

11. Gibbs KW, Semler MW, Driver BE, et al. Noninvasive Ventilation for Preoxygenation during Emergency Intubation. *N Engl J Med*. 2024;390(23):2165-2177.

12. Casey JD, Janz DR, Russell DW, et al. Bag-Mask Ventilation during Tracheal Intubation of Critically Ill Adults. *N Engl J Med*. 2019;380(9):811-821.

13. Russell DW, Casey JD, Gibbs KW, et al. Effect of Fluid Bolus Administration on Cardiovascular Collapse Among Critically Ill Patients Undergoing Tracheal Intubation: A Randomized Clinical Trial. *JAMA*. 2022;328(3):270-279.

14. Knapp S, Kofler J, Stoiser B, et al. The assessment of four different methods to verify

tracheal tube placement in the critical care setting. *Anesth Analg*. 1999;88(4):766-770.

15. Vukmir RB, Heller MB, Stein KL. Confirmation of endotracheal tube placement: a miniaturized infrared qualitative CO2 detector. *Ann Emerg Med*. 1991;20(7):726-729.

16. Grmec S, Mally S. Prehospital determination of tracheal tube placement in severe head injury. *Emerg Med J*. 2004;21(4):518-520.

17. Grmec S. Comparison of three different methods to confirm tracheal tube placement in emergency intubation. *Intensive Care Med*. 2002;28(6):701-704.

18. Takeda T, Tanigawa K, Tanaka H, Hayashi Y, Goto E, Tanaka K. The assessment of three methods to verify tracheal tube placement in the emergency setting. *Resuscitation*. 2003;56(2):153-157.

19. Ornato JP, Shipley JB, Racht EM, et al. Multicenter study of a portable, hand-size, colorimetric end-tidal carbon dioxide detection device. *Ann Emerg Med*. 1992;21(5):518-523.

20. Varon AJ, Morrina J, Civetta JM. Clinical utility of a colorimetric end-tidal CO2 detector in cardiopulmonary resuscitation and emergency intubation. *J Clin Monit*. 1991;7(4):289-293.

21. Rabitsch W, Nikolic A, Schellongowski P, et al. Evaluation of an end-tidal portable ETCO2 colorimetric breath indicator (COLIBRI). *Am J Emerg Med*. 2004;22(1):4-9.

22. Hayden SR, Sciammarella J, Viccellio P, Thode H, Delagi R. Colorimetric end-tidal CO2 detector for verification of endotracheal tube placement in out-of-hospital cardiac arrest. *Acad Emerg Med*. 1995;2(6):499-502.

23. Linko K, Paloheimo M, Tammisto T. Capnography for detection of accidental oesophageal intubation. *Acta Anaesthesiol Scand*. 1983;27(3):199-202.

24. Puntervoll SA, Søreide E, Jacewicz W, Bjelland E. Rapid detection of oesophageal intubation: take care when using colorimetric capnometry. *Acta Anaesthesiol Scand*. 2002;46(4):455-457.

25. Driver B, Law J. Confirmation of correct endotracheal tube placement in adults. UpToDate. Accessed June 3, 2026. https://www.uptodate.com/contents/confirmation-of-correct-endotracheal-tube-placement-in-adults

26. Asai T, Shingu K. Case report: A normal capnogram despite esophageal intubation. *Can J Anaesthesia-journal Can D Anesthesie*. 2001;48:1025-1028.

27. MacLeod BA, Heller MB, Gerard J, Yealy DM, Menegazzi JJ. Verification of endotracheal tube placement with colorimetric end-tidal CO2 detection. *Ann Emerg Med*. 1991;20(3):267-270.

28. Driver BE, Prekker ME, Casey JD. Use of a Bougie vs Endotracheal Tube With Stylet and Successful Intubation on the First Attempt Among Critically Ill Patients Undergoing Tracheal Intubation-Reply. *JAMA*. 2022;327(15):1503-1504.

29. Topjian AA, Raymond TT, Atkins D, et al. Part 4: Pediatric basic and advanced life support 2020 American heart association guidelines for cardiopulmonary resuscitation and

emergency cardiovascular care. *Pediatrics*. 2021;147(Supplement 1):e2020038505D.

30. Honardar MR, Posner KL, Domino KB. Delayed Detection of Esophageal Intubation in Anesthesia Malpractice Claims: Brief Report of a Case Series. *Anesth Analg*. 2017;125(6):1948-1951.

31. Lee D. Inquest into the death of Emiliana Obusan. 2021. Accessed March 20, 2026. https://coroners.nsw.gov.au/documents/findings/2021/OBUSAN_Emiliana_-_Findings.pdf

32. English C. Amended version of Finding into Death Without Inquest regarding XH. 2021. Accessed March 20, 2026. https://www.coronerscourt.vic.gov.au/sites/default/files/COR%202019%2001998%20XH%20Finding%20Amended.pdf

33. Schaller RJ, Huff JS, Zahn A. Comparison of a colorimetric end-tidal CO2 detector and an esophageal aspiration device for verifying endotracheal tube placement in the prehospital setting: a six-month experience. *Prehosp Disaster Med*. 1997;12(1):57-63.

34. Knapp S, Kofler J, Stoiser B, et al. The assessment of four different methods to verify tracheal tube placement in the critical care setting. *Surv Anesthesiol*. 2000;44(1):12-13.

35. Silvestri S, Ladde JG, Brown JF, et al. Endotracheal tube placement confirmation: 100% sensitivity and specificity with sustained four-phase capnographic waveforms in a cadaveric experimental model. *Resuscitation*. 2017;115:192-198.

36. Sayah AJ, Peacock WF, Overton DT. End-tidal CO2 measurement in the detection of esophageal intubation during cardiac arrest. *Surv Anesthesiol*. 1991;XXXV(2):78.

37. Bozeman WP, Hexter D, Liang HK, Kelen GD. Esophageal detector device versus detection of end-tidal carbon dioxide level in emergency intubation. *Ann Emerg Med*. 1996;27(5):595-599.

38. Heidegger T. Management of the Difficult Airway. *N Engl J Med*. 2021;384(19):1836-1847.

39. Pfuntner A, Wier LM, Stocks C. Most frequent procedures performed in US hospitals, 2011: statistical brief# 165. In: *Healthcare Cost and Utilization Project (HCUP) Statistical Briefs.* (http://www.ncbi.nlm.nih.gov/ books/NBK174682/). Agency for Healthcare Research and Quality; 2013.

40. Cairns C, Kang K. National Hospital Ambulatory Medical Care Survey: 2021 emergency department summary tables. https://www.cdc.gov/nchs/data/nhamcs/web_tables/2021-nhamcs-ed-web-tables-508.pdf

41. Schandelmaier S, Briel M, Varadhan R, et al. Development of the Instrument to assess the Credibility of Effect Modification Analyses (ICEMAN) in randomized controlled trials and meta-analyses. *CMAJ*. 2020;192(32):E901-E906.
